# Mapping renal impairment and cardiac structure and function: a comprehensive analysis of prospective cohort study, Mendelian randomization and shared genetic etiology

**DOI:** 10.1101/2025.08.24.25333620

**Authors:** Haozhang Huang, Jin Liu, Xiaozhao Lu, Shiqun Chen, Yang Zhou, Jiyan Chen, Ning Tan, Wei Jiang, Yong Liu

## Abstract

**Background:** The elevated risk of heart failure in kidney dysfunction is well-identified. However, the influence of early alteration of renal impairment on cardiac structure and function, as well as their shared genetic susceptibility associations have not been well reported.

**Aim:** To investigate the associations between renal function and cardiac parameters using cardiac magnetic resonance images (CMR).

**Methods:** Of 29,546 European subjects underwent CMR test were recruited from UK Biobank study. Multivariable generalized linear regression was used to analyze the association of different kidney function indexes with CMR-traits. Genetic correlation between early renal impairment and CMR-traits were evaluated by linkage disequilibrium score regression and Mendelian randomization.

**Design:** A comprehensive analysis of prospective cohort study, Mendelian randomization and shared genetic etiology.

**Results:** Observational and genetic correlation analyses consistently reveal that a mild reduction in estimated glomerular filtration rate (eGFR) is independently associated with lower biventricular volume parameters and left ventricular cardiac output (LVCO), respectively. Through a rigorous application of Heritability Estimation from and Summary Statistics (ρ-HESS) and cross-trait meta-analysis, we identified 20 novel shared loci (e.g. rs2472297, located at *CYP1A1*) between eGFR based on cystatin C decline and reduced right ventricular end systolic volume (RVESV), and 2 loci (rs4371638, located at *SHROOM3*; rs34591452, located at *STRA6*) showed strong evidence of colocalization (probability for H4>0.95). Pathway enrichment analysis revealed that pathways of eGFR decline and reduced RVESV associated with enzyme inhibitor activity, endopeptidase regulator activity, and cysteine−type endopeptidase inhibitor activity.

**Conclusions:** Our study indicates significant observational and genetic correlations between early renal impairment and lower biventricular volume parameters and LVCO.

## Introduction

Chronic kidney disease (CKD) poses a significant global health burden, affecting approximately 10% of the population (1). About half of CKD patients succumb to cardiovascular disease, particularly heart failure (HF), without progressing to end-stage renal disease (2, 3). Substantial evidence shows that severe kidney dysfunction, as measured by estimated glomerular filtration rate (eGFR) and urine albumin-to-creatinine ratio (uACR), is independently associated with heart failure and adverse cardiac structure and function as evaluated by echocardiography (4–7).

Cardiac magnetic resonance (CMR) imaging is considered the gold standard for the comprehensive assessment of cardiac morphology and function, particularly for the right ventricle, due to its superior image quality and reproducibility (8). Comprehensive mapping of cardiac structure and function in individuals with mild to moderate kidney dysfunction remains insufficiently explored.

The UK Biobank (UKB), one of the largest multimodal studies, provides high-quality standardized CMR examinations and dense genotype data. This resource enables the establishment of genome-wide association studies (GWAS) profiles for CMR phenotypes, which can aid in evaluating genetic susceptibility to disease, developing new drug targets, and offering genetic insights into the kidney-heart connection (9).

Hence, this study aims to examine: 1) observational associations between renal function (CKD, eGFR [serum creatinine, cystatin C], uACR) and cardiac morphology, function, and geometry; 2) genetic associations between renal function and CMR measurements using linkage disequilibrium score regression (LDSC) and Mendelian randomization (MR); and 3) shared risk single nucleotide polymorphisms (SNPs) by employing Heritability Estimation from Summary Statistics (ρ-HESS), cross-trait meta-analyses, and colocalization (Central illustration).

## Materials and methods

### Participants

From the UKB dataset (n = 502,355), we excluded participants lacking information on race, ethnicity, or valid kidney function measurements (serum creatinine [SCr], cystatin C, uACR). Among the remaining individuals with kidney function measurements (n = 451,433), 29,546 participated in the ongoing CMR study, which includes comprehensive measurements of the four chambers and aorta (Figure 1). Detailed ethical approval are available in the supplementary materials. This study received ethical approval for UKB research from the NHS National Research Ethics Service on June 17, 2011, with renewal on June 18, 2021 (ref 11/NW/0382).). All participants provided written informed consent.

**Figure. 1.**
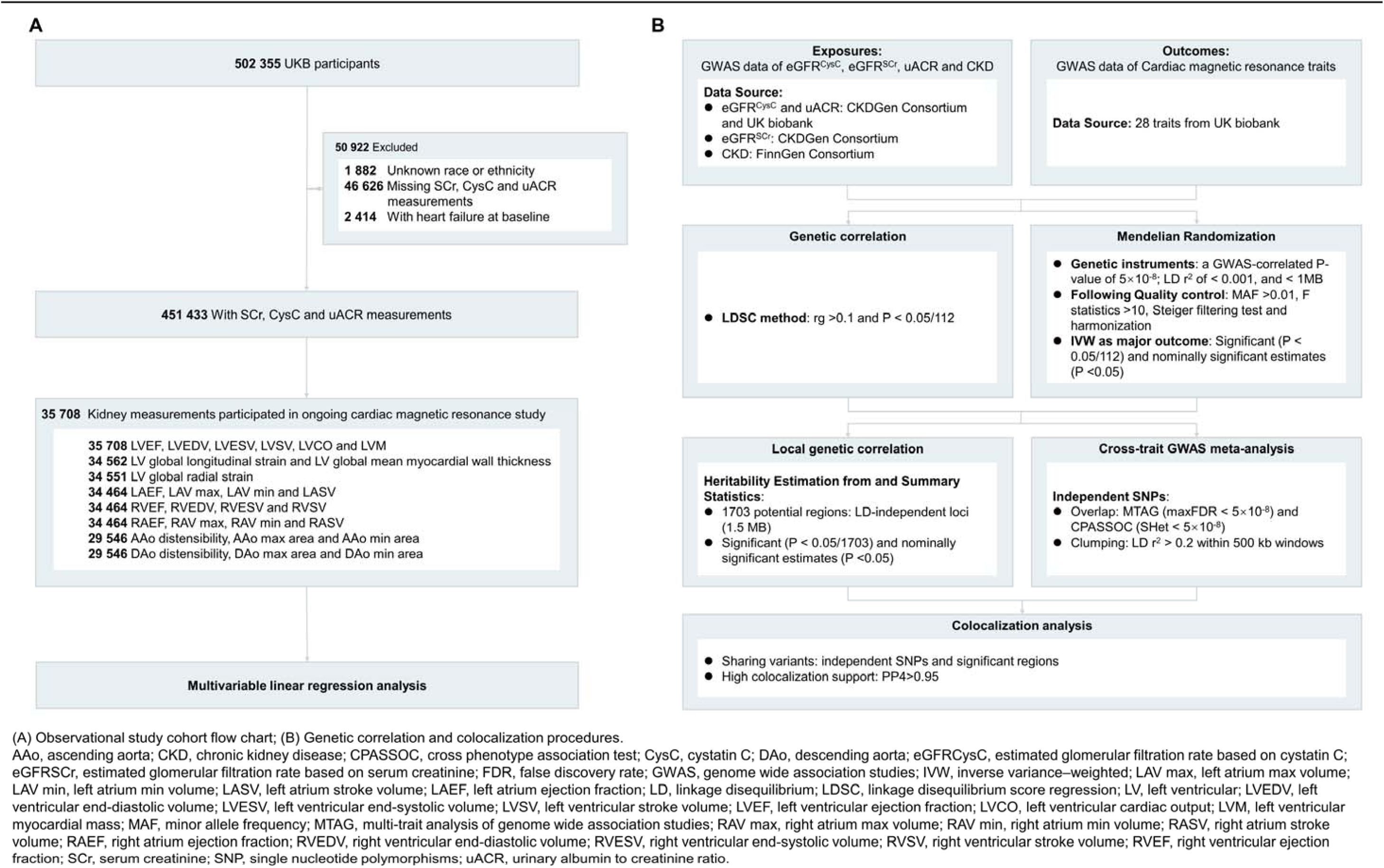
Main design of this study

### Derivation of CMR Parameters and Arterial Stiffness

We generated 28 CMR traits from the raw CMR images (UKB Data-Field 100003) based on short-axis, long-axis, and aortic cine images. These traits include 10 left ventricle traits, 4 left atrium traits, 4 right ventricle traits, 4 right atrium traits, and 3 traits each for the ascending and descending aorta. The left ventricle traits encompass volumetric measures such as left ventricular end-diastolic volume (LVEDV), end-systolic volume (LVESV), stroke volume (LVSV), ejection fraction (LVEF), cardiac output (LVCO), and myocardial mass (LVM), along with global measures for myocardial wall thickness at end-diastole and various strain metrics. Right ventricle traits include RVEDV, RVESV, stroke volume (RVSV), and ejection fraction (RVEF). For the atria, we derived maximum and minimum volumes, stroke volumes, and ejection fractions. For the aorta, we assessed maximum and minimum areas, along with distensibility for both the ascending and descending aorta. Detailed protocols and analysis methods have been published previously (10–13). Detailed Kidney Function are available in the supplementary materials.

### Statistical Analysis Observational Epidemiology

Descriptive statistics are presented as means with standard deviations for continuous variables and counts with percentages for categorical variables. Categorical variables were compared using chi-square tests, while normally distributed continuous variables were compared using Student’s t-test. Multivariable linear regression models adjusted for age, sex, race, smoking status, body mass index (BMI), systolic blood pressure (SBP), heart rate (HR), hypertension medication use, coronary artery disease (CAD), and diabetes mellitus (DM) were employed to evaluate associations between kidney function and CMR measures. A significant threshold of P < 0.05 was used, with all P-values derived from two-tailed hypothesis tests.

### Heritability and Genetic Correlation

We employed precomputed linkage disequilibrium (LD) scores from the 1000 Genomes Project, focusing on SNPs in the HapMap 3 SNP set. SNPs not consistent with the reference panel (minor allele frequency [MAF] ≤ 0.01 or INFO score ≤ 0.9) were excluded. LDSC was applied to estimate heritability for individual traits and assess genetic correlations between kidney function and cardiac measures, utilizing GWAS summary statistics and LD scores from European ancestry reference data.

### MR Analysis

To explore potential causality and directionality between kidney function and CMR traits, we conducted MR analyses using large-scale GWAS datasets. Summary-level data for eGFR based on CysC (eGFR^CysC^) and uACR were obtained from a meta-analysis of European-ancestry participants from the CKDGen Consortium and UKB (14). The meta-analysis of the GWAS of eGFR based on creatine (eGFR^SCr^) was from CKDGen Consortium (15). The meta-analysis for eGFR based on creatinine (eGFRSCr) was sourced from CKDGen. GWAS summary statistics for CKD comprised 10,039 cases and 396,706 controls from the FinnGen study, identified via ICD codes. Summary-level data related to cardiac structure and function were obtained from the UKB meta-analysis (9).

We identified causal relationships using genetic instruments that reflect different aspects of renal pathophysiology: 1) index SNPs for eGFR^CysC^ (Table S1), 2) eGFRSCr (Table S2), 3) CKD based on ICD (Table S3), and 4) uACR (Table S4). Genetic instruments were selected based on a GWAS-correlated P-value of 5 × 10^-8^, LD r² < 0.001, and being located within 1 MB of the index variant. Quality control filters applied included: MAF > 0.01; an F-statistic for each instrument (F = [(N-k-1)/k] * [R²/(1-R²)]) of < 10 indicating weak instruments; the Steiger filtering test to avoid reverse causality; and harmonization of exposure and outcome data.

For each analysis, we used the inverse variance-weighted (IVW) MR method while accounting for random effects. We also reported the P-value for the intercept from MR Egger regression to check for horizontal pleiotropy. Sensitivity analyses included the weighted median method and MR-PRESSO to address measurement errors and selection bias. A combination of approaches provides robust evidence for potential causal relationships (16–18).

All analyses were performed using the TwoSampleMR (version 0.4.25) and MRPRESSO (version 1.0) packages in R (version 4.3). With 112 MR estimates (4 × 28), a Bonferroni-corrected P-value threshold was set at 0.05/112 (4.46 × 10^-4^), with P < 0.05 deemed nominally significant.

### Bidirectional and Multivariable MR

We first conducted bidirectional MR analyses to estimate causal effects of kidney function decline on significant cardiac measurements. Subsequently, we performed multivariable MR (MVMR) analyses to estimate direct effects of each kidney function measure, accounting for genetic correlations between eGFR, CKD, and uACR (19).

### Shared Genetic Correlation Analysis

We utilized the ρ-HESS method to estimate local SNP heritability and genetic correlation, analyzing shared genetic etiology and causal effects between kidney function and cardiac measurements. In a cohort of European ancestry individuals, we identified 1,703 approximately LD-independent regions, averaging 1.5 MB in size. Local SNP heritability and genetic correlations were calculated using the 1000 Genomes Project as a reference, with Bonferroni correction applied for multiple testing. Shared risk SNPs were detected using multi-trait analysis of GWAS (MTAG) and cross-phenotype association test (CPASSOC), with colocalization analysis performed via coloc to evaluate shared causal variants. Detailed methods are available in the supplementary materials.

### GO and Pathway Enrichment Analysis

Gene Ontology (GO) and Kyoto Encyclopedia of Genes and Genomes (KEGG) pathway analyses for differentially expressed genes (DEGs) were conducted using the Database for Annotation, Visualization, and Integrated Discovery (version 6.8) (20). A cutoff criterion of more than 2 enriched genes and P < 0.05 was used.

## Results

### Baseline Characteristics

Among the 29,546 participants included in the analysis, the mean age was 54.9 years (SD = 7.4), with 51.1% being women. Baseline characteristics stratified by eGFR^CysC^ categories are summarized in Table 1. Normal renal function (eGFR^CysC^ > 90 mL/min/1.73m²) was observed in 17,482 participants (59.1%), while 11,598 (39.3%) had mild renal dysfunction (60 ≤ eGFR^CysC^ < 90 mL/min/1.73m²) and 466 (1.6%) had moderate to severe renal dysfunction (eGFR^CysC^ < 60 mL/min/1.73m²). Participants in the moderate or severe renal dysfunction group tended to be older and predominantly male, exhibiting a more adverse cardiovascular risk profile, including higher proportions of previous smoking, elevated BMI, DM, HF, CAD, and stroke, as well as increased likelihood of receiving hypertension and lipid treatment. In cardiac four-chamber measurements, lower biventricular volumes (LVEDV, LVESV, LVSV, RVEDV, RVESV, and RVSV), elevated LA volume, thicker LV walls (LVM and global mean myocardial wall thickness), and higher LV global longitudinal strain were found in individuals with lower eGFR^CysC^. For aortic measurements, decreased aortic distensibility and increased aortic area were observed in the severe renal dysfunction group. Additional baseline characteristics stratified by eGFR^SCr^, uACR, and CKD are presented in Supplement 1 (Tables S5-S7).

**Table 1.** Study cohort characteristics.

| Total, No. | Overall<br>29 546 | eGFR<sup>CysC</sup>, ml/min/1.73m<sup>2</sup> |  |  | **P** |
| --- | --- | --- | --- | --- | --- |
|  |  | ≥90<br>17 482 | 60-89<br>11 598 | <60<br>466 |  |
| Age, years | 54.9 (7.4) | 52.5 (7.1) | 58.2 (6.5) | 61.5 (5.6) | <0.001 |
| Female, No. (%) | 15103 (51.1) | 9577 (54.8) | 5318 (45.9) | 208 (44.6) | <0.001 |
| Race and ethnicity, No. (%) |  |  |  |  | <0.001 |
| White | 28730 (97.2) | 16956 (97.0) | 11319 (97.6) | 455 (97.6) |  |
| Black | 169 (0.6) | 135 (0.8) | 33 (0.3) | 1 (0.2) |  |
| Asian | 302 (1.0) | 156 (0.9) | 139 (1.2) | 7 (1.5) |  |
| Other/mixed | 345 (1.2) | 235 (1.3) | 107 (0.9) | 3 (0.6) |  |
| Smoking Status, No. (%) |  |  |  |  | <0.001 |
| Current | 1812 (6.1) | 929 (5.3) | 850 (7.3) | 33 (7.1) |  |
| Previous | 9790 (33.2) | 5550 (31.8) | 4054 (35.1) | 186 (40.0) |  |
| Never | 17884 (60.7) | 10976 (62.9) | 6662 (57.6) | 246 (52.9) |  |
| BMI, kg/m<sup>2</sup> | 26.4 (4.1) | 25.7 (3.8) | 27.5 (4.3) | 29.0 (5.3) | <0.001 |
| SBP, mmHg | 136.9 (18.7) | 134.6 (18.3) | 140.1 (18.8) | 140.6 (18.9) | <0.001 |
| DBP, mmHg | 81.4 (10.4) | 80.5 (10.3) | 82.6 (10.4) | 81.8 (10.2) | <0.001 |
| HR, bpm | 62.5 (10.3) | 62.3 (10.1) | 62.8 (10.5) | 63.8 (11.3) | <0.001 |
| DM, No. (%) | 832 (2.8) | 400 (2.3) | 389 (3.4) | 43 (9.2) | <0.001 |
| CAD, No. (%) | 835 (2.8) | 347 (2.0) | 447 (3.9) | 41 (8.8) | <0.001 |
| Stroke, No. (%) | 232 (0.8) | 96 (0.5) | 122 (1.1) | 14 (3.0) | <0.001 |
| AMI, No. (%) | 330 (1.1) | 124 (0.7) | 188 (1.6) | 18 (3.9) | <0.001 |
| AF, No. (%) | 298 (1.0) | 112 (0.6) | 171 (1.5) | 15 (3.2) | <0.001 |
| SCr, mg/dL | 0.8 (0.2) | 0.8 (0.1) | 0.9 (0.2) | 1.1 (0.3) | <0.001 |
| CysC, mg/dL | 0.9 (0.1) | 0.8 (0.1) | 1.0 (0.1) | 1.3 (0.2) | <0.001 |
| uACR, mg/g | 9.4 [5.9, 15.8] | 10.0 [6.2, 16.6] | 8.7 [5.6, 14.7] | 8.7 [5.8, 17.1] | <0.001 |
| Hypertension treatment, No. (%) | 2445 (8.3) | 945 (5.4) | 1362 (11.7) | 138 (29.6) | <0.001 |
| Lipid treatment, No. (%) | 3099 (10.5) | 1351 (7.7) | 1599 (13.8) | 149 (32.0) | <0.001 |
| AAo distensibility, 10<sup>-3</sup>/mmHg | 1.8 (1.3) | 2.0 (1.4) | 1.6 (1.1) | 1.3 (0.9) | <0.001 |
| AAo max area, mm<sup>2</sup> | 853.1 (190.1) | 836.0 (187.9) | 876.8 (189.9) | 907.3 (206.7) | <0.001 |
| AAo min area, mm<sup>2</sup> | 776.0 (185.3) | 755.6 (183.2) | 804.2 (183.7) | 838.6 (199.0) | <0.001 |
| DAo distensibility, 10<sup>-3</sup>/mmHg | 2.4 (1.5) | 2.6 (1.6) | 2.1 (1.3) | 1.8 (1.0) | <0.001 |
| DAo max area, mm<sup>2</sup> | 475.5 (97.3) | 464.0 (93.8) | 491.5 (99.5) | 506.7 (110.5) | <0.001 |
| DAo min area, mm<sup>2</sup> | 419.0 (92.1) | 405.8 (88.5) | 437.3 (93.4) | 455.3 (103.0) | <0.001 |
| LAEF, % | 61.2 (9.3) | 61.9 (8.5) | 60.4 (10.1) | 58.4 (12.2) | <0.001 |
| LAV max, mL | 72.6 (23.2) | 72.2 (22.0) | 73.3 (24.8) | 74.0 (26.7) | <0.001 |
| LAV min, mL | 29.3 (15.3) | 28.4 (13.5) | 30.4 (17.2) | 32.8 (21.9) | <0.001 |
| LASV, mL | 43.4 (11.8) | 43.8 (11.6) | 42.9 (12.0) | 41.2 (11.5) | <0.001 |
| LVCO, L/min | 5.4 (1.3) | 5.4 (1.3) | 5.4 (1.2) | 5.3 (1.3) | 0.019 |
| LV global circumferential strain, % | -22.3 (3.4) | -22.3 (3.2) | -22.2 (3.5) | -21.9 (4.0) | 0.001 |
| LVEF, % | 59.6 (6.1) | 59.6 (5.8) | 59.5 (6.3) | 59.3 (7.0) | 0.456 |
| LVEDV, mL | 148.2 (33.7) | 149.0 (33.6) | 147.3 (33.7) | 143.8 (35.5) | <0.001 |
| LVESV, mL | 60.6 (19.3) | 60.8 (18.9) | 60.3 (19.6) | 59.3 (21.8) | 0.023 |
| LV global longitudinal strain, % | -18.5 (2.8) | -18.5 (2.7) | -18.4 (2.9) | -18.2 (3.3) | 0.008 |
| LV global mean myocardial wall thickness, mm | 5.7 (0.8) | 5.6 (0.8) | 5.8 (0.8) | 5.9 (0.8) | <0.001 |
| LVM, g | 86.3 (22.2) | 85.2 (22.2) | 87.9 (22.1) | 88.1 (22.5) | <0.001 |
| LVSV, mL | 87.7 (19.1) | 88.2 (19.1) | 87.0 (19.2) | 84.4 (19.5) | <0.001 |
| LV global radial strain, % | 45.1 (8.3) | 44.9 (8.0) | 45.2 (8.6) | 45.1 (9.7) | 0.006 |
| RAEF, % | 47.1 (9.3) | 47.3 (9.0) | 46.8 (9.7) | 46.9 (10.6) | 0.001 |
| RAV max | 86.7 (27.5) | 87.2 (26.8) | 86.0 (28.3) | 84.1 (28.7) | <0.001 |
| RAV min | 46.4 (18.9) | 46.4 (18.2) | 46.4 (20.0) | 45.6 (21.6) | 0.6569 |
| RASV, mL | 40.3 (13.3) | 40.7 (13.1) | 39.6 (13.5) | 38.5 (13.5) | <0.001 |
| RVEF, % | 57.3 (6.1) | 57.3 (5.9) | 57.2 (6.3) | 57.3 (7.1) | 0.4223 |
| RVEDV, mL | 157.1 (37.1) | 158.1 (37.5) | 156.0 (36.6) | 149.4 (34.9) | <0.001 |
| RVESV, mL | 67.8 (21.1) | 68.2 (21.4) | 67.3 (20.8) | 64.5 (20.6) | <0.001 |
| RVSV, mL | 89.3 (20.3) | 89.9 (20.1) | 88.6 (20.4) | 85.0 (20.2) | <0.001 |

Abbreviations: AAo, ascending aorta; AF, atrial fibrillation; AMI, acute myocardial infarction; BMI, body mass index; CAD, coronary artery disease; CHOL, Cholesterol; CysC, cystatin C; DAo, descending aorta; DBP, diastolic blood pressure; DM, diabetes mellitus; eGFR<sup>CysC</sup>, estimated glomerular filtration rate based on cystatin C; eGFR<sup>SCr</sup>, estimated glomerular filtration rate based on serum creatinine; HDL-C, high density lipoprotein cholesterol; HR, heart rate; LDL, low density lipoprotein cholesterol; LAV max, left atrium max volume; LAVmin, left atrium min volume; LASV, left atrium stroke volume; LAEF, left atrium ejection fraction; LV, left ventricular; LVEDV, left ventricular end-diastolic volume; LVESV, left ventricular end-systolic volume; LVSV, left ventricular stroke volume; LVEF, left ventricular ejection fraction; LVCO, left ventricular cardiac output; LVM, left ventricular myocardial mass; RAV max, right atrium max volume; RAV min, right atrium min volume; RASV, right atrium stroke volume; RAEF, right atrium ejection fraction; RVEDV, right ventricular end-diastolic volume; RVESV, right ventricular end-systolic volume; RVSV, right ventricular stroke volume; RVEF, right ventricular ejection fraction; SBP, systolic blood pressure; SCr: serum creatinine; uACR, urinary albumin to creatinine ratio.

### Observational Associations with Kidney Function and Cardiac measurements

In multivariable adjusted models, a significant positive association was observed between eGFR^CysC^ (per 1 SD decrease) and biventricular volume and aortic area (all P ≤ 0.001; Table 2 and Figure 2). In addition, lower level of eGFR^CysC^ was associated with less LVM (β =-0.66; 95% CI,-0.85 to-0.47; P <0.001), lower LAEF (β =-0.28; 95% CI,-0.41 to-0.16; P<0.001) and lower RAEF (β =-0.14; 95% CI,-0.26 to-0.01; P = 0.032). In terms of eGFR^SCr^, consistent associations were observed in biventricular parameters and DAo area. Similarly, positive associations were observed between CKD and biventricular volume as well as LVM. In addition, higher level of uACR was associated with larger AAo area and right atrial parameters. Other results were shown in Supplement 1 (Table S8).

**Figure. 2.**
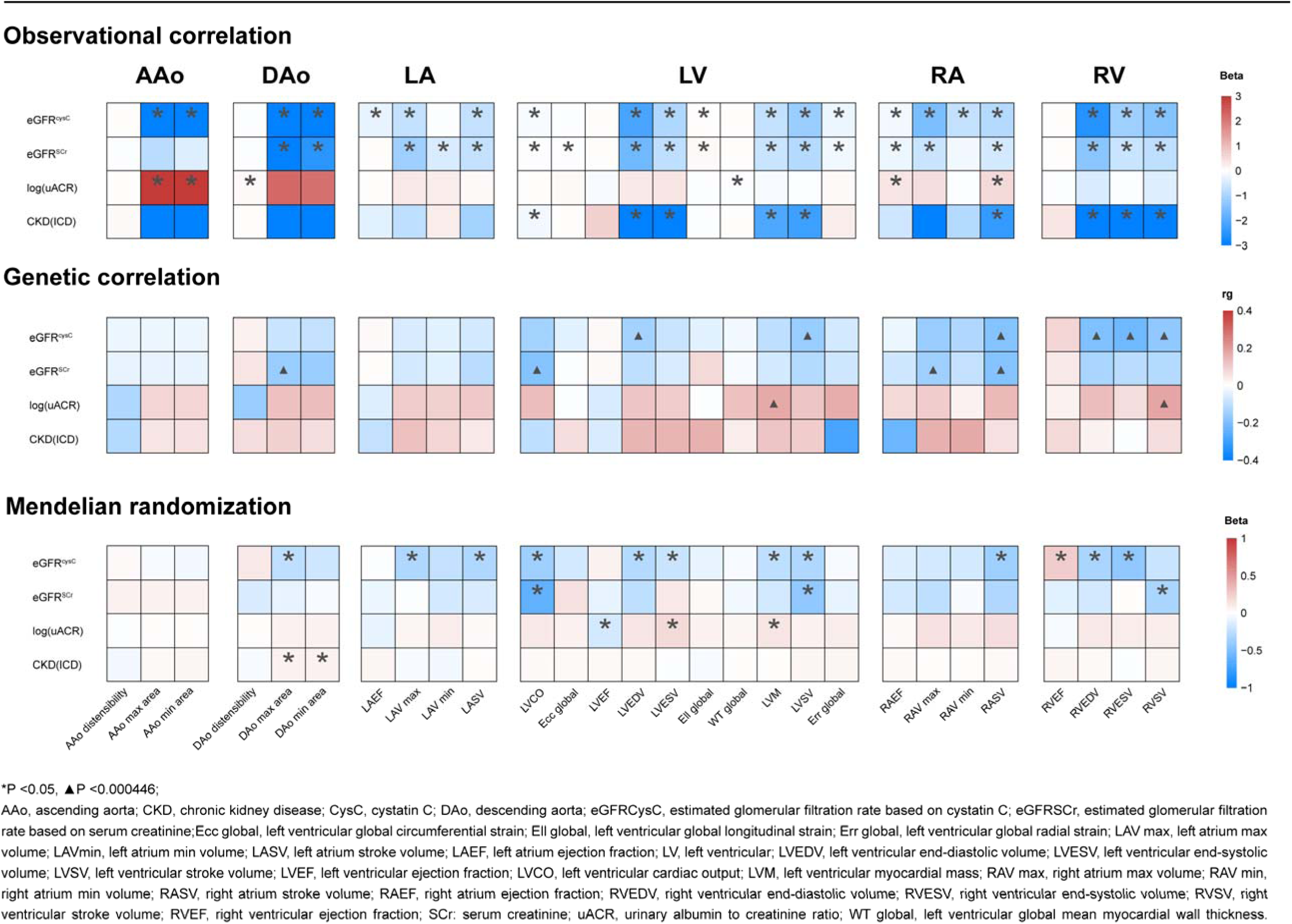
Association Between kidney function decline and cardiac Imaging Traits

**Table 2.**
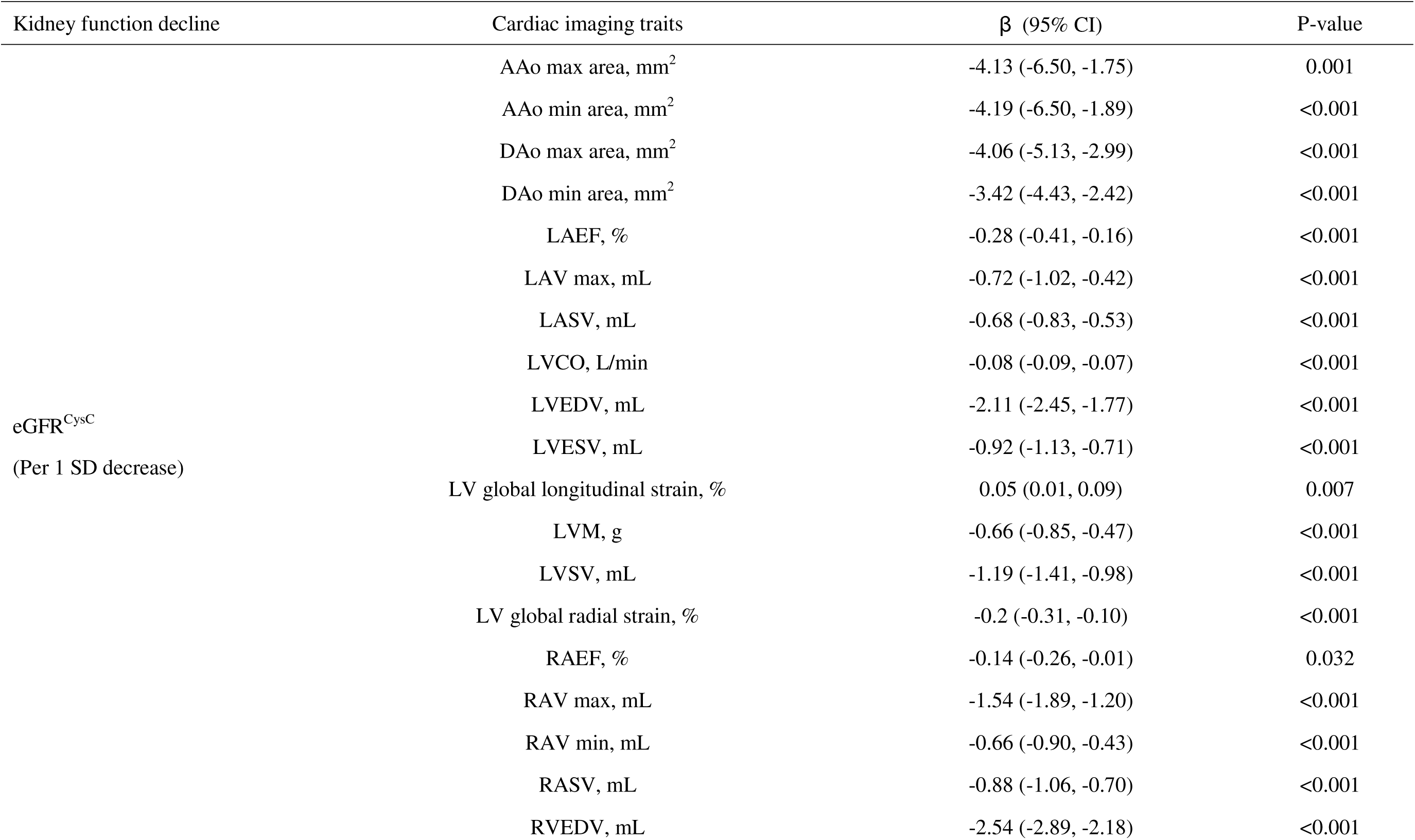
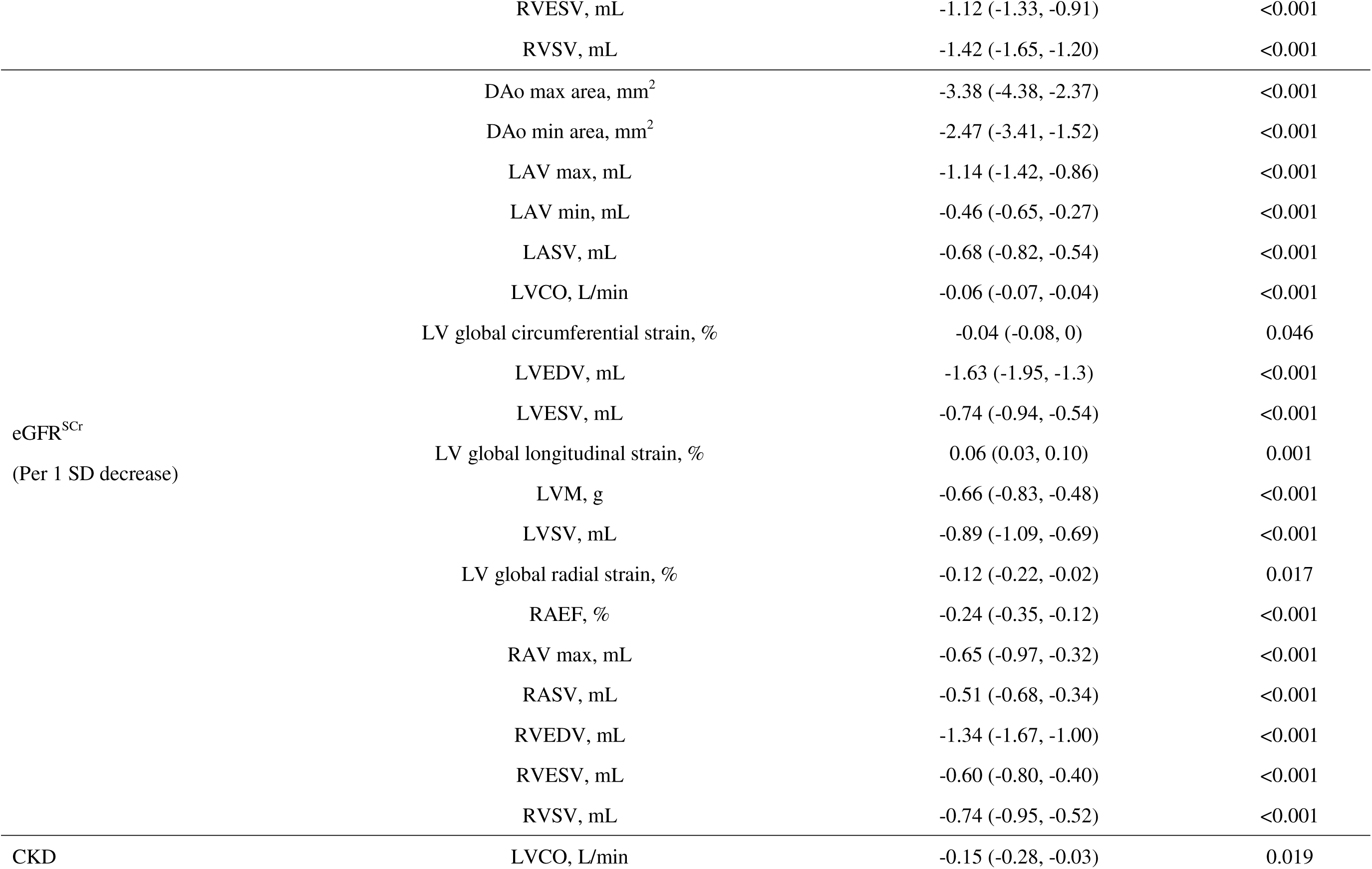
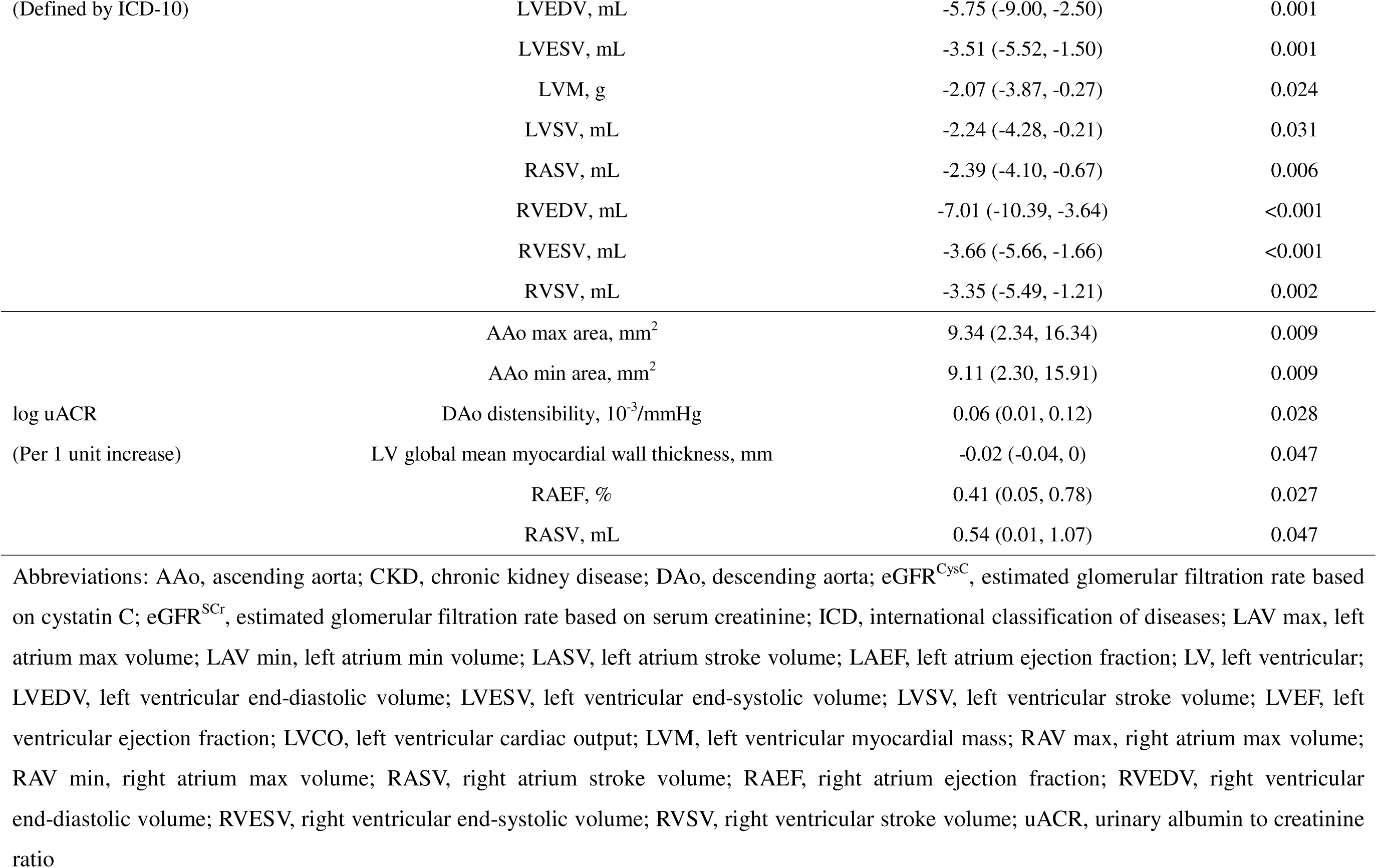
Significant Multivariable regression results for kidney function decline and cardiac imaging traits.

### MR Analysis

We conducted a comprehensive MR study from genetically predicted eGFR, CKD, and uACR on 28 cardiac structure and function, and identified 20 significant or nominal significant outcomes influenced by kidney function decline. The fixed-effects IVW method was used as our principal MR analytic approach (Figure 2).

In Table 3, genetically instrumented reduced log-eGFR^CysC^ was found to decrease RVESV (β =-0.43 mL, 95% CI:-0.53 mm to-0.33 mm, P = 1.60×10^-5^) and RVEDV (β =-0.34 mL, 95% CI:-0.43 mL to-0.25 mL, P = 0.0004). Heterogeneity was not observed with a Cochran Q-derived P value > 0.05 and the P value for the MR-Egger intercept is > 0.05 (Table S9). Similarly, Weighted median, MR-PRESSO, c-MRL, GSMR method also showed that reduced log-eGFR^CysC^ was significantly correlated with an increased risk of RVESV and RVEDV (P < 0.05) (Table S10 and Figure S1). Reverse causation was not observed in genetically determined reduced l log-eGFR^CysC^ with RVESV and RVEDV (Table S10). In multivariable MR analysis controlling for CKD and uACR, we found consistent evidence for a causal independent effect of genetically predicted reduced log-eGFR^CysC^ on RVESV and RVEDV (P < 0.05; Table S11). No outliers were identified with MR-PRESSSO and the leave-one-out plot as well as funnel plots (Figure S2).

**Table 3.**
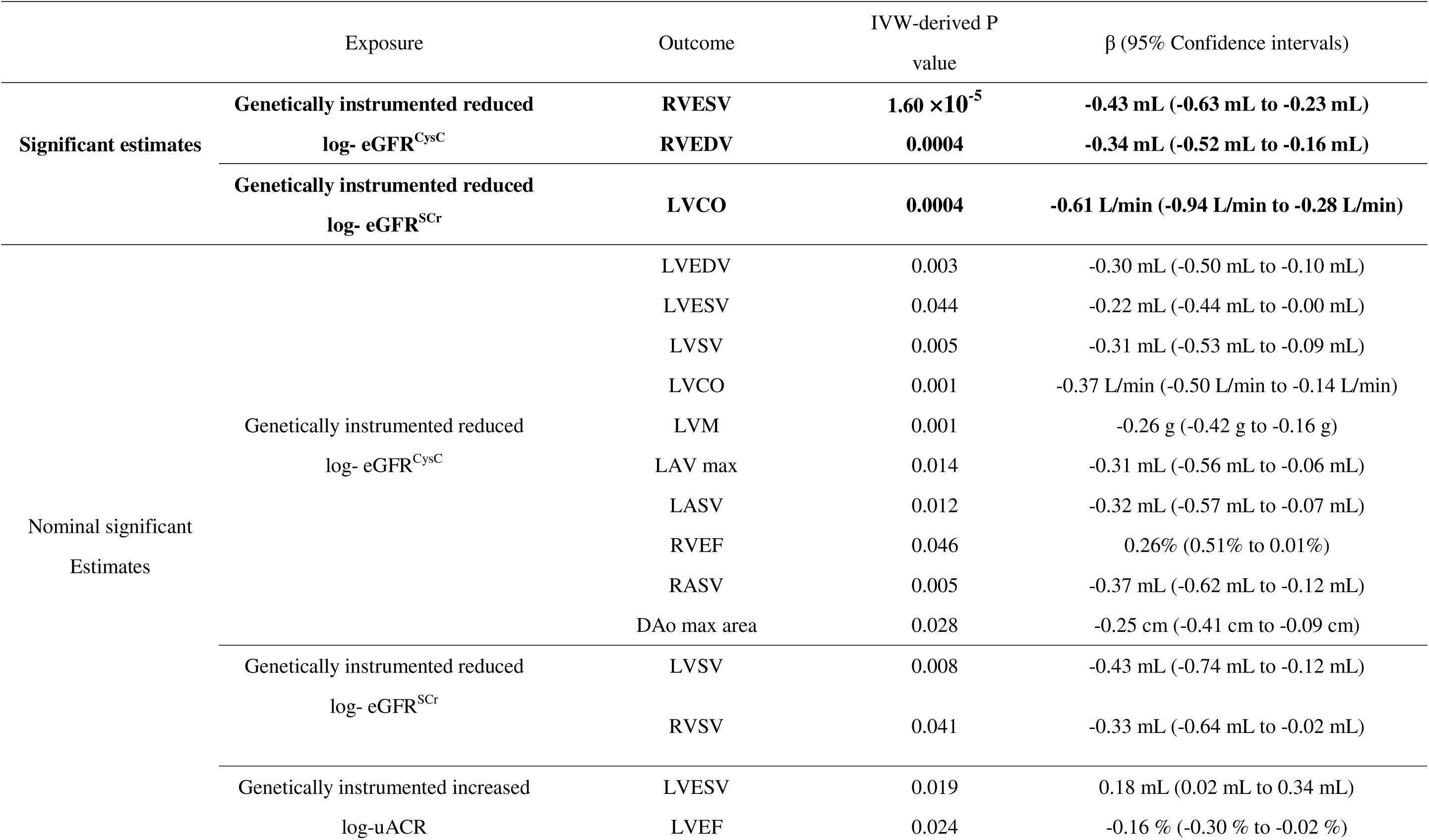
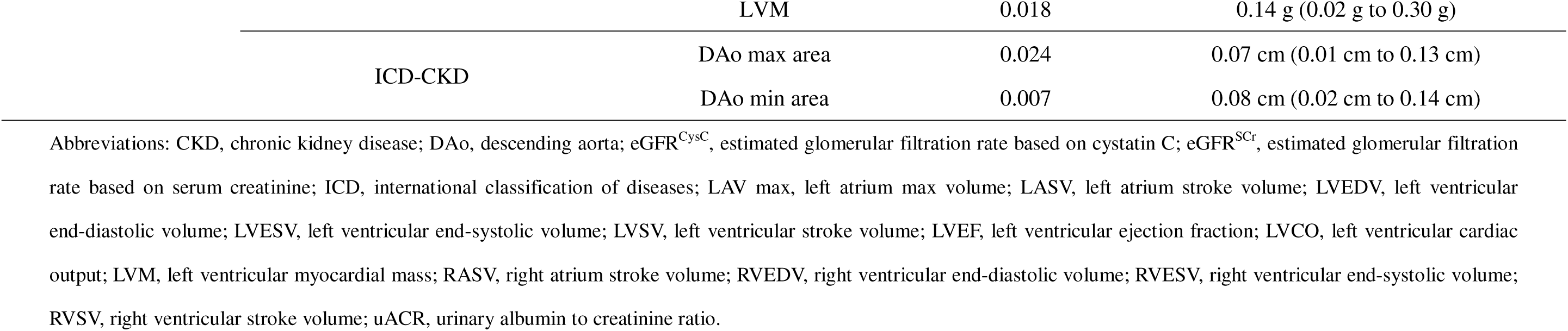
Significant causal estimates of kidney function decline and cardiac Imaging Traits.

Genetically instrumented reduced log-eGFR^SCr^ was found to decrease LVCO (β =-0.61 L/min, 95% CI:-0.88 L/min to-0.44 L/min, P = 0.0004). MR-PRESSO, c-MRL, GSMR method also showed that reduced log-eGFR^SCr^ was significantly correlated with an increased risk of left ventricular cardiac output (P < 0.05) (Table S10 and Figure S3). Sensitivity analysis of causal association between eGFR^SCr^ and LVCO was shown in Supplement 1 (Figure S4). Our findings indicate a nominal significant correlation between the CKD and the increasing DAo max area (β = 0.07 cm, 95% CI: 0.04 cm to 0.10 cm, P = 0.024), also observed in DAo min area (β = 0.08cm, 95% CI: 0.05 cm to 0.11 cm, P = 0.0007). Genetically instrumented increased uACR was found to increase LVESV, LVM, and LV regional radial strain (P < 0.05) (Table S10 and Figures S5-S6).

In summary, there were 17 suggestive cardiac structures and functions, including LVEDV, LVESV, LVSV, LVCO, LVM, LV regional peak circumferential strain, LVEF, LAV max, LASV, RVEDV, RVEF, RASV, aorta maximum area, aorta minimum area, potentially influenced by kidney function (P < 0.05). Details are presented in Supplement 1 (Table S12).

### Dissecting Shared Genetic Etiology

We applied bivariate LDSC to estimate the genetic correlation (without constrained intercept) between kidney function and cardiac measurements (Table S13), focusing on traits with overlapping causal relationships identified by MR and genetic associations discovered via LDSC (Figure 2). The genetic correlation between eGFR^CysC^ and RVESV was found to be 0.22 (P < 0.001, LDSC; Table S14). The liability-scale SNP heritability estimates were 11.8% for eGFR^CysC^ and 20.3% for RVESV. After constraining the LDSC intercept under the assumption of no sample overlap, the genetic correlation remained significant but slightly weakened (P < 0.001). The genetic correlation between eGFR^SCr^ and LVCO was 0.19 (P < 0.001), while the correlation between uACR and LVM was similarly 0.19, and between eGFRSCr and LVCO it was 0.18 (P < 0.001).

### Identification of Shared SNPs for Kidney Function and Cardiac Measurements

To enhance our power in identifying shared genetic SNPs, we performed GWAS meta-analysis between eGFR and RVESV due to the strong genetic relationships. A total of 105 genome-wide significant independent SNPs (P < 5 × 10^-8^) were revealed in both MTAG and CPASSOC analyses, including 99 newly identified shared SNPs (Tables S15-S16). The results from MTAG were consistent with those from CPASSOC, suggesting reliability and minimal bias in the MTAG assumptions. Local genetic correlations were estimated, revealing two significant regions (P < 0.05/number of regions, ρ-HESS) (Figure 3, Table S17, and Figure S7). A total of 20 risk-independent SNPs showed consistent significance when examined by ρ-HESS with P < 0.05 (Tables S18-S19), with the most significant shared SNP being rs2472297 (PCPASSOC = 2.86 × 10^-33^) located at *CYP1A1*.

**Figure. 3.**
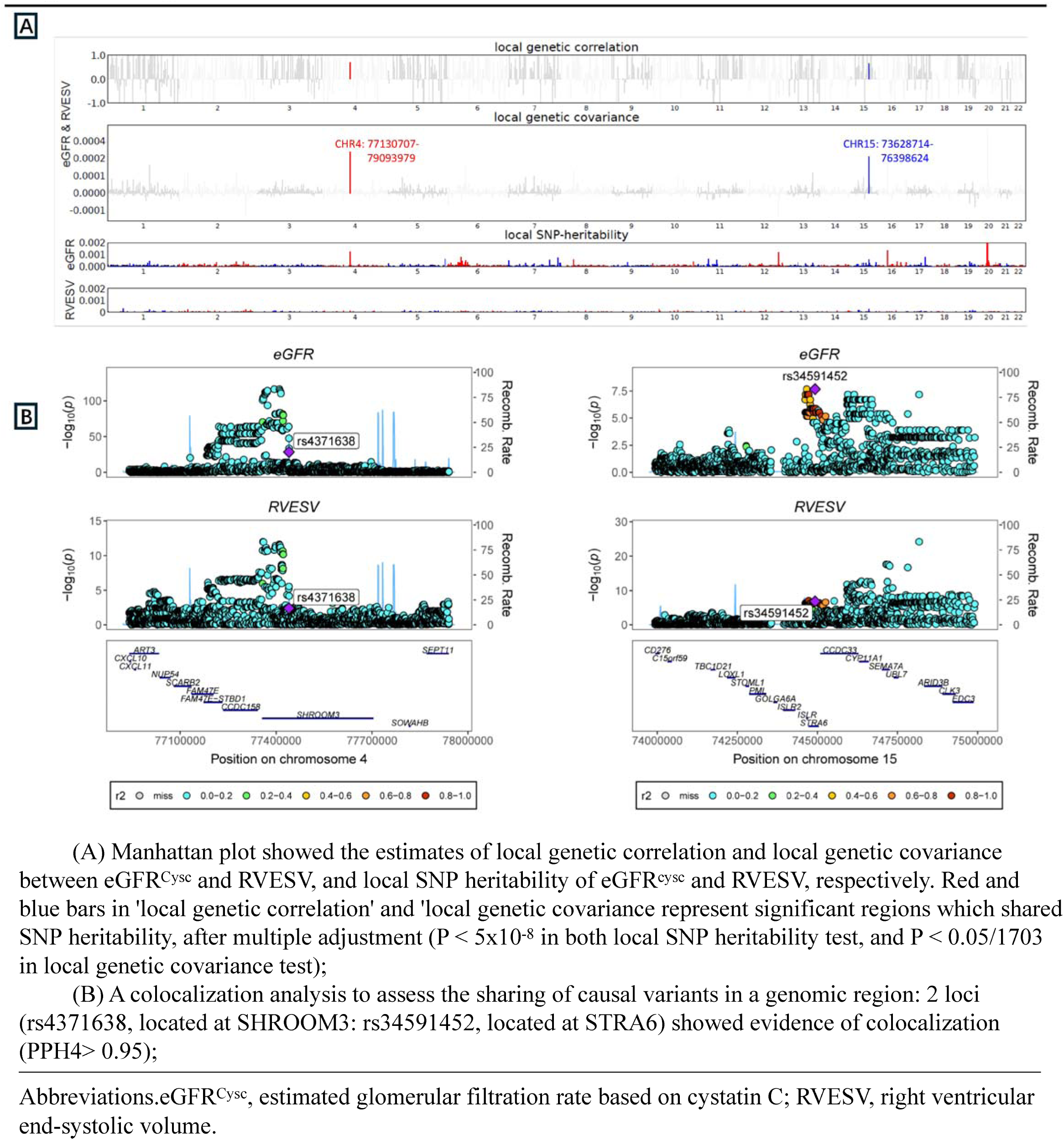
Local genetic correlation between eCiFR^cYsc^ and RVESV

For the analysis between eGFR^SCr^ and LVCO, a total of 67 significant independent SNPs were identified (Tables S20-S21) and two regions were pinpointed (P < 0.05, Table S22 and Figure S8). Additionally, ρ-HESS and cross-trait meta-analysis identified three novel loci (rs10066035 at *NSD1*, rs3812036 at *SLC34A1*, and rs9905761 at *BCAS3*) shared between eGFRSCr decline and reduced LVCO (Table S23). For the relationship between uACR and LVM, a total of three significant independent SNPs were identified (Tables S24-S25), but no regions were defined (P < 0.05, Figure S9).

### Colocalization Analysis

Colocalization analysis was performed to determine whether the genetic variants driving the associations in the two traits are shared or distinct. Shared loci between eGFR^CysC^ and RVESV colocalized at the same candidate causal SNPs (PPH4 > 0.95). Among the 20 novel pleiotropic loci identified between eGFR^CysC^ decline and reduced RVESV, two loci (rs4371638 at *SHROOM3* and rs34591452 at *STRA6*) exhibited significant colocalization (PPH4 > 0.95) (Figure 3 and Figure S10).

### Pathway Enrichment Analysis

The pathway enrichment analysis revealed that the 99 newly identified shared SNPs of eGFR and RVESV were significantly enriched in four GO terms or KEGG pathways, including those related to enzyme inhibitor activity, endopeptidase regulator activity, and cysteine-type endopeptidase inhibitor activity, with enriched genes being > 2 and P_FDR < 0.05 (Figure S12).

## Discussion

This study represents the first extensive investigation into the potential causal effects of early kidney impairment on a broad spectrum of cardiac structure and function assessed through CMR traits. The observational and genetic data presented emphasize the importance of declining kidney function as a critical risk factor for alterations in cardiac structure and function. Notably, genetically instrumented early reductions in eGFR were significantly associated with decreased biventricular volume parameters and LVCO. Furthermore, we identified 20 novel shared loci associated with eGFR^CysC^ decline and reduced RVESV, with two loci (rs4371638 at *SHROOM3* and rs34591452 at *STRA6*) demonstrating strong evidence of colocalization.

CMR is the gold standard for evaluating cardiac structure and function, allowing precise measurement of diastolic and systolic volumes and enabling the calculation of ejection fractions. Additionally, CMR flow studies facilitate the estimation of forward flow through semilunar and atrioventricular (AV) valves, leading to accurate computations of regurgitant fractions and cardiac output. The UKB provides high-quality standardized CMR examinations alongside extensive genotype data, which previous studies have utilized alongside deep learning techniques to explore cardiac phenotypes more accurately. Given the challenges in longitudinally tracking cardiac structure and function using CMR, we opted for a cross-sectional analysis to examine correlations between kidney function and cardiac structure, complemented by MR analysis to investigate potential causal relationships.

Many large observational cohort studies, particularly those focused on CKD, have reported that declining renal function correlates with worsening diastolic function, a phenomenon confirmed by animal studies (4, 21–24). For instance, Buckley et al. reported that lower eGFR was linked to significant increases in left ventricular end-diastolic volume index and deteriorating diastolic measures among adults free of HF at baseline, while uACR did not correlate with changes in cardiac structure or function (23). Conversely, Cai et al. found that more advanced CKD at baseline correlated with increases in LVM and volume, as well as greater diastolic dysfunction (24). However, these results should be interpreted cautiously, as they may not entirely apply to community populations. Although up to 40% of CKD patients exhibit left ventricular hypertrophy (25), of interest, several studies report no independent association between lower kidney function and LVM. This discrepancy may arise from restrictive enrollment criteria in those studies (26, 27), limiting the variability of cardiac structure. Some research suggests a positive relationship between high GFR and LV hypertrophy, implying a potential link between hyperfiltration and LV hypertrophy (28, 29). In mildly reduced eGFR stages, traditional risk factors such as hypertension may explain associations with LVM. Indeed, in community-based cohorts, the connection between low eGFR and LV hypertrophy was only significant before controlling for confounders (30). Consequently, the relationship between eGFR and left ventricular function and structure, especially within normal or mildly reduced ranges, may be complex. Buckley et al. found kidney function decline was not significantly associated with right ventricular fractional area change (23), although the limitations of cardiac ultrasound do not detract from the importance of assessing the right heart structure in relation to kidney function decline (31). Our study, conducted within a healthier community cohort, suggests that early cardiac changes relating to renal function decline differ in healthy populations. Mild renal dysfunction may correlate more closely with reduced stroke volume and decreased effective circulating blood volume.

The underlying mechanisms may include several factors. In the UKB population, 59.1% of participants had normal renal function (eGFR > 90 mL/min/1.73m²), while 39.3% had mild renal dysfunction (60 ≤ eGFR < 90 mL/min/1.73m²) and 2.7% had moderate to severe dysfunction (eGFR < 60 mL/min/1.73m²). In contrast, an Atherosclerosis Risk in Communities study indicated that 36.5% had reduced eGFR < 60 mL/min/1.73m² (4).

Previous studies utilizing echocardiography to assess left and right ventricular function have highlighted preload, afterload, contractility, and lusitropy as key determinants. Pathophysiological mechanisms of RV failure may involve acute or chronic load abnormalities or myocardial dysfunction coexisting in clinical RV failure states (32). Increased afterload results in RV hypertrophy, and although compensatory mechanisms initially maintain stroke volume, increased heart rate exacerbates myocardial oxygen demand while reducing filling time, leading to decreased cardiac output. In patients with CKD, sympathetic nervous system (SNS) dysfunction, characterized by chronic SNS activation, can lead to renal vasoconstriction and reduced renal blood flow, impairing kidney function over time. Increased SNS activity is common in CKD patients, even in early stages, suggesting a significant role in renal disease pathophysiology (33, 34) In generally healthy populations, even minor declines in kidney function may trigger early cardiac compensatory mechanisms. Our findings indicate that reduced cardiac output can be linked to diminished end-diastolic volume, reflecting a decrease in effective circulating blood volume. Renal impairment may stem from circulatory dysfunction, including splanchnic arterial vasodilation and reduced effective arterial blood volume. Patients with impaired renal function often receive diuretics to manage symptoms, which can further reduce effective circulating volume mediated by venous dilation and increased urinary output (35). Although we considered relevant confounding factors (hypertension, diabetes, coronary artery disease), renal dysfunction is nonetheless associated with cardiovascular structural changes, predisposing patients to complications. Thus, the relationship between gradual kidney function decline, stroke volume reduction, and effective circulating blood volume is complex, reflecting an early stage of cardiac compensation. Future investigations should encompass thorough evaluations of renal function alongside cardiac structural changes, especially in individuals considered relatively healthy. Comprehensive assessments can deepen our understanding of the underlying mechanisms connecting kidney and heart health.

In our study, we screened 9 SNPs with pleiotropic effects in the cross-trait meta-analyses and ρ-HESS of eGFR^CysC^ and RVESV. Previous studies showed that *SHROOM3* (rs4371638) associated with CKD showed associations with both the high levels of oxidatively damaged DNA and genomic instability (36, 37).

*SEMA7A*(rs11854025) encodes a member of the semaphorin family of proteins. The encoded protein is found on activated lymphocytes and erythrocytes and may be involved in immunomodulatory and neuronal processes. *SEMA7A* is increased in patients with acute aortic dissection and is a Novel Biomarker in Kidney Renal Clear Cell Carcinoma (38, 39). *NRG4* (rs10851885) is associated with imbalance in glucose metabolism and obesity, and significantly lower in patients with end-stage kidney disease (40–42). Notably, the independent shared SNP was rs2472297 (*P*_CPASSOC_ = 2.86×10^-33^) located at *CYP1A1*. The cytochrome P450 proteins are monooxygenases which catalyze many reactions involved in drug metabolism and synthesis of cholesterol, steroids, and other lipids. Previous studies have linked *CYP1A1* to ischemic stroke and coronary heart disease (43–45). These could be some potential targets for intervention.

In summary, our findings provide crucial insights for future research. Our logistic regression analyses established significant links between renal function deterioration and cardiac structure, demonstrating a notable association between reduced eGFR and both cardiac output and biventricular volume in the general population. We further confirmed a causal relationship between declining kidney function and alterations in cardiac structure and function, identifying key genes associated with early compensatory mechanisms. We recommend early CMR assessments for patients with mild renal insufficiency to accurately evaluate right ventricular function.

## Limitations

Several limitations should be acknowledged. Firstly, there existed considerable variation in the estimations of causal effects derived from various variants (eGFR, CKD, uACR, Tables S1-S4). Nonetheless, other potential sources of heterogeneity might exist, though they do not necessarily introduce bias. Secondly, it is crucial to acknowledge that UKB comprises a community cohort primarily of European descent; consequently, the findings may vary among distinct populations or ethnic backgrounds. The occurrence of severe renal dysfunction is notably low in this population who underwent CMR testing, thus limiting the study’s ability to generalize its findings beyond the effects of mild renal dysfunction on cardiac function and structure within the general community. Additionally, the absence of quantitative assessments of myocardial fibrosis using late gadolinium enhancement-MRI imaging represents another noteworthy limitation.

## Conclusion

In conclusion, this comprehensive analysis reveals both observational and genetic associations between early renal impairment and cardiac structure and function assessed through CMR traits. Our findings demonstrate that early declines in kidney function significantly correlate with reduced biventricular volume and LVCO. Further research is needed to elucidate the precise biological mechanisms underlying these associations.

## Supplemental Materia

Tables S1-S29 Figures S1-S12

## Supporting information

checklist

## Data Availability

All data produced in the present study are available upon reasonable request to the authors

## Acknowledgements

We thank the blogger (orange_milk_sugar, Wenyan Chen) for guidance on methods.

## Author contributions

Dr Yong Liu had full access to all the data in the study and takes responsibility for the integrity of the data and the accuracy of the data analysis. Drs Jin Liu, Haozhang Huang, and Xiaozhao Lu contributed equally to this article. Concept and design: Wei Jiang and Yong Liu. Acquisition, analysis, or interpretation of data: Jin Liu and Yong Liu. Drafting of the manuscript: Haozhang Huang, Xiaozhao Lu, and Yang Zhou. Statistical analysis: Jin Liu and Haozhang Huang. Obtained funding: Yong Liu and Shiqun Chen.

Administrative, technical, or material support: Ning Tan and Jiyan Chen. Supervision: Shiqun Chen, Wei Jiang and Yong Liu.

## Data availability

This study utilized data from the UK Biobank Resource (project number 99231). GWAS summary statistics for the 82 heart imaging traits are freely available for download from Zenodo: https://zenodo.org/record/7239166. Additional GWAS summary statistics and information from published projects of the CKDGen Consortium can be accessed at: https://ckdgen.imbi.uni-freiburg.de/.

---

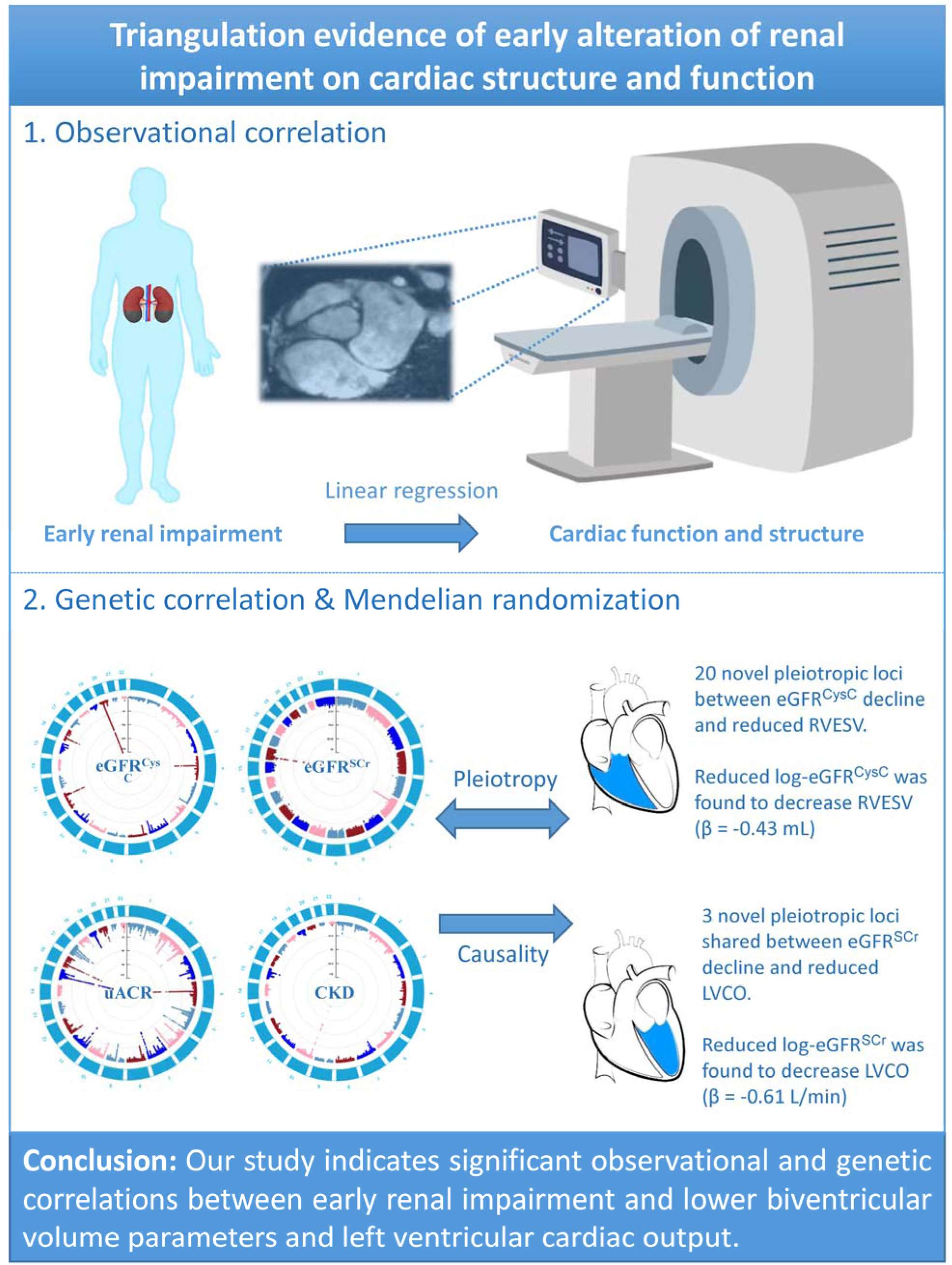

