## Supplement Method and Figure for "Mapping renal impairment and cardiac structure and function: a comprehensive analysis of prospective cohort study, Mendelian randomization and shared genetic etiology"

**Supplement Legends**

**Figure S1.** GSMR of causal association between eGFR^CysC^ and RVESV.

**Figure S2.** Sensitivity analysis of causal association between eGFR^CysC^ and RVESV.

**Figure S3.** GSMR of causal association between eGFR^SCr^ and LVCO.

**Figure S4.** Sensitivity analysis of causal association between eGFR^SCr^ and LVCO.

**Figure S5.** GSMR of causal association between uACR and LVM.

**Figure S6.** Sensitivity analysis of causal association between uACR and LVM.

**Figure S7.** Local genetic covariance estimates.

**Figure S8.** Local genetic correlation between eGFR^Scr^ and LVCO.

**Figure S9.** Local genetic correlation between uACR and LVM.

**Figure S10.** Colocalization analysis of eGFR^CysC^ and RVESV.

**Figure S11.** Study design of Mendelian randomization analysis.

**Figure S12.** Pathway enrichment analysis of shared SNPs of eGFR and RVESV.

**Table S1**. 192 index SNPs represented genetically predicted eGFR^CysC^.

**Table S2.** 201 index SNPs represented genetically predicted eGFR^SCr^.

**Table S3**. 5 index SNPs represented genetically predicted CKD.

**Table S4.** 57 index SNPs represented genetically predicted uACR.

**Table S5.** Study cohort characteristics stratified by uACR level.

**Table S6.** Study cohort characteristics stratified by eGFR^SCr^.

**Table S7.** Study cohort characteristics stratified by CKD.

**Table S8.** Multivariable linear associations between renal function decline and cardiac traits.

**Table S9.** Sensitivity analysis of significant causal estimates of kidney function decline and cardiac Imaging Traits.

**Table S10.** Bidirectional Mendelian randomization estimates for the causal effect of genetically predicted reduced eGFR and significant cardiac measurements.

**Table S11.** Multivariable Mendelian randomization estimates for the causal effect of kidney function decline and significant cardiac measurements.

**Table S12.** Causal estimates of kidney function decline and cardiac imaging traits.

**Table S13.** Genetic correlations between kidney function and cardiac traits.

**Table S14.** Genetic correlations between eGFR and cardiac traits.

**Table S15**. Independent Significant pleiotropic SNPs identified by cross-trait meta-analysis of eGFR^CysC^ and RVESV.

**Table S16.** Independent novel significant pleiotropic SNPs identified by cross-trait meta-analysis of eGFR^CysC^ and RVESV.

**Table S17**. Significant regions identified by ρ-HESS of eGFR^CysC^ and RVESV.

**Table S18.** Significant pleiotropic SNPs identified by cross-trait meta-analysis and ρ-HESS (p < 0.05) of eGFR^CysC^ and RVESV.

**Table S19**. Significant pleiotropic SNPs identified by cross-trait meta-analysis and ρ-HESS (p < 0.05/1703) of eGFR^CysC^ and RVESV.

**Table S20.** Independent significant pleiotropic SNPs identified by cross-trait meta-analysis of eGFR^SCr^ and LVCO.

**Table S21.** Independent novel significant pleiotropic SNPs identified by cross-trait meta-analysis of eGFR^SCr^ and LVCO.

**Table S22.** Significant regions identified by ρ-HESS (p < 0.05) of eGFR^SCr^ and LVCO.

**Table S23.** Significant pleiotropic SNPs identified by cross-trait meta-analysis and ρ-HESS of eGFR^SCr^ and LVCO.

**Table S24**. Independent Significant pleiotropic SNPs identified by cross-trait meta-analysis of uACR and LVM.

**Table S25.** Independent novel significant pleiotropic SNPs identified by cross-trait meta-analysis of uACR and LVM.

**Table S26.** Multivariable linear associations between renal function decline and cardiac traits after excluding patients with current smoking.

**Table S27.** Multivariable linear associations between renal function decline and cardiac traits in patients with complete information of covariates.

**Table S28.** Multivariable linear associations between renal function decline and cardiac traits with additional adjustment for specific antihypertensive drugs use (ACEI/ARB, CCB, β-blockers).

**Table S29.** Results from multivariable Mendelian randomization analysis.

**Supplement Method**

**Kidney Function**

We extracted serum cystatin C (CysC) and serum creatinine (SCr) from blood biochemistry under the UKB Data-Field 30720 and 30700. CysC levels were determined by Immuno-turbidimetric (Siemens plc, Siemens Advia 1800). SCr levels were determined by enzymatic (Beckman Coulter [UK], Ltd, Beckman Coulter AU5800). The Chronic Kidney Disease Epidemiology Collaboration formula was used to calculate the SCr- and CysC-based eGFR (1, 2). History of CKD was according to International Classification of Diseases 9 (ICD-9 Code: 585) or 10 (ICD-10 Code: I12-13, N18), end stage renal disease (ESRD) and Dialysis. Urinary albumin values below the detection limit of the used assays were set to the lower limit of detection, and the uACR was assessed in mg/g and calculated as urinary albumin (mg/L)/urinary creatinine (mg/dL) × 100.

**Observational Epidemiology**

Various sensitivity analyses will be performed in the linear regression analysis. First, we will exclude patients who are still smoking, considering the potential impact of smoking on both kidney function and cardiac structure. Second, we will perform on individuals with complete information on the covariates. Finally, we will use multiple models for thorough adjustment: we will further adjust for various medications, including beta-blockers and RASS blockers (Tables S26-S28).

**Mendelian randomization design**

Large-scale genome-wide association studies conducted over the last decade have uncovered numerous genetic variants associated with cardiometabolic traits and risk factors. These discoveries have enabled the Mendelian randomization (MR) design, which uses genetic variation as a natural experiment to improve causal inferences from observational data. MR studies, an IV-based method to infer the causality between intermediate phenotypes and disease, have been widely conducted in cardiometabolic research (3, 4). Multivariable Mendelian Randomization Analysis (MVMR) MVMR is an extension of MR that takes into account the multiplicity between traits. In this study, we used MVMR to assess whether an outcome (CMR-related traits) was related to exposure (kidney function) as well as “independent” causality between kidney function, hypertension, coronary heart disease, diabetes (Figure S11 and Table S29).

**Local Genetic Correlation Analysis**

ρ-HESS is a method to estimate local SNP-heritability and genetic correlation. We estimated the local genetic correlations to examine the significant overlapping traits of shared genetic etiology and causal effect between kidney function and cardiac measurements at the local independent region in the genome using ρ-HESS(5, 6). In human genome with European ancestry, there were 1703 potential regions that were approximately LD-independent loci, with an average size of nearly 1.5 MB. Then we calculated the local SNP heritability for two traits and the genetic correlation between two traits using the 1000 Genomes Project as the reference provided on the ρ-HESS webpage. Nominal significant was defined as 0.05 and Bonferroni correction was then applied to adjust for multiple testing (0.05/ the number of regions).

**Cross-trait GWAS Meta-analysis**

To detect the shared risk SNPs in the significant overlapping traits of shared genetic etiology and causal effect between kidney function and cardiac measurements, we performed two cross-trait meta-analyses, including multi-trait analysis of GWAS (MTAG) and cross phenotype association test (CPASSOC)(7, 8). MTAG is a generalized meta-analysis method that enhances statistical power to estimate the genotypic and phenotypic variance-covariance matrices to generate trait-specific estimates for each SNP. SNPs were restricted with MAF ≥ 0.01 and sample size N ≥ (2/3) × 90th percentile. MTAG adjusts for possible errors by using bivariate LD score regression when sample overlap is present. MTAG is suitable when all variants have the same effect sizes on traits and generate trait-specific association statistics. We calculated the upper bound for the false discovery rate (‘maxFDR’) to examine the assumptions on the equal variance covariance. In addition, as a sensitivity analysis, CPASSOC integrates association evidence from multiple traits’ GWAS summary statistics when the variant is correlated to at least one trait. We utilized the SHet version to assume heterogeneous effects across traits. Subsequently, independent SNPs were obtained (were most significantly associated with the phenotype) using the PLINK "clumping" function through applying the following parameters: –clump-p1 5e-8 –clump-p2 1e-5 –clump-r2 0.2 –clump-kb 500. Novel loci were defined as independent SNPs without linkage disequilibrium (LD r^2^ > 0.2 within 1000-kb windows) with significant SNPs in the original single-trait GWAS. We prioritized independent SNPs that were genome-wide significant (P < 5×10^-8)^ in the cross-trait meta-analyses using both MTAG and CPASSOC and were in significant regions identified by ρ-HESS.

**Colocalization Analysis**

We next performed a colocalization analysis through coloc(9). coloc uses a Bayesian algorithm to generate posterior probabilities for five mutually exclusive hypotheses regarding the sharing of causal variants in a genomic region, namely H0 (no association), H1 or H2 (association to one trait only), H3 (association to both traits, two distinct SNPs), and H4 (association to both traits, one shared SNP). We extracted summary statistics for variants within 1.0 Mb of the index SNP at each shared locus and calculated the posterior probability for H4 (PPH4). A locus was considered colocalized if PPH4 greater than 0.95.

**Figure S1.** GSMR of causal association between eGFR^CysC^ and RVESV.

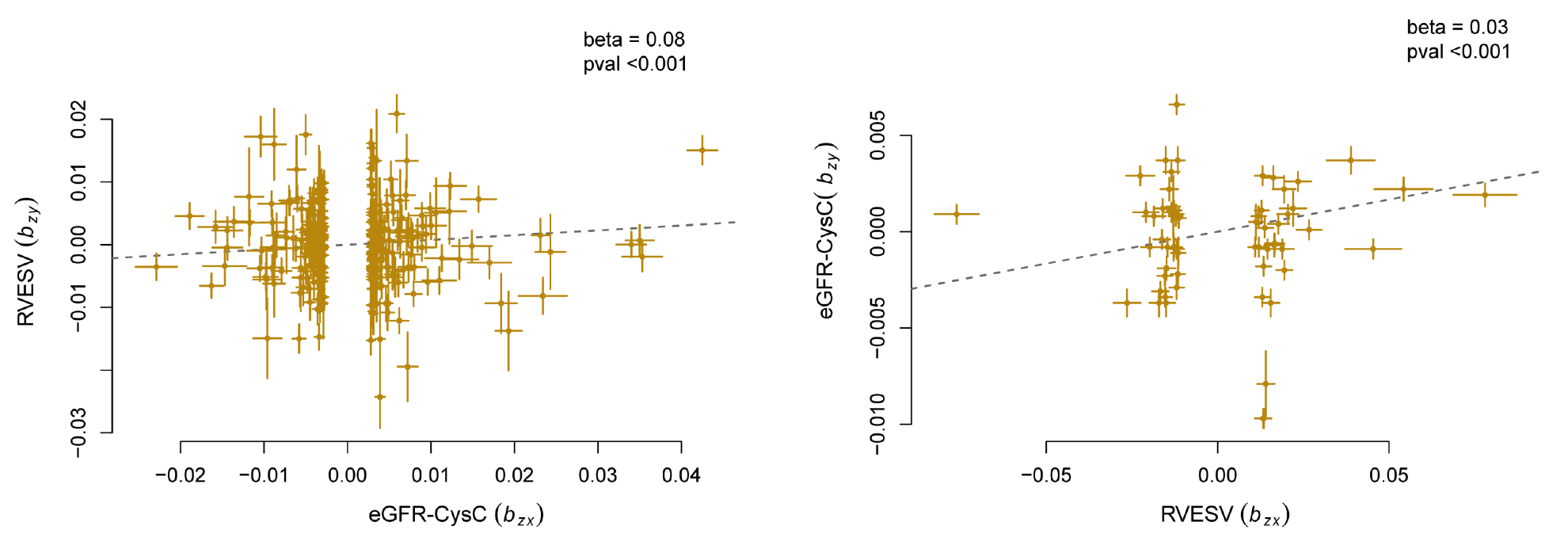

**Figure S2.** Sensitivity analysis of causal association between eGFR^CysC^ and RVESV.

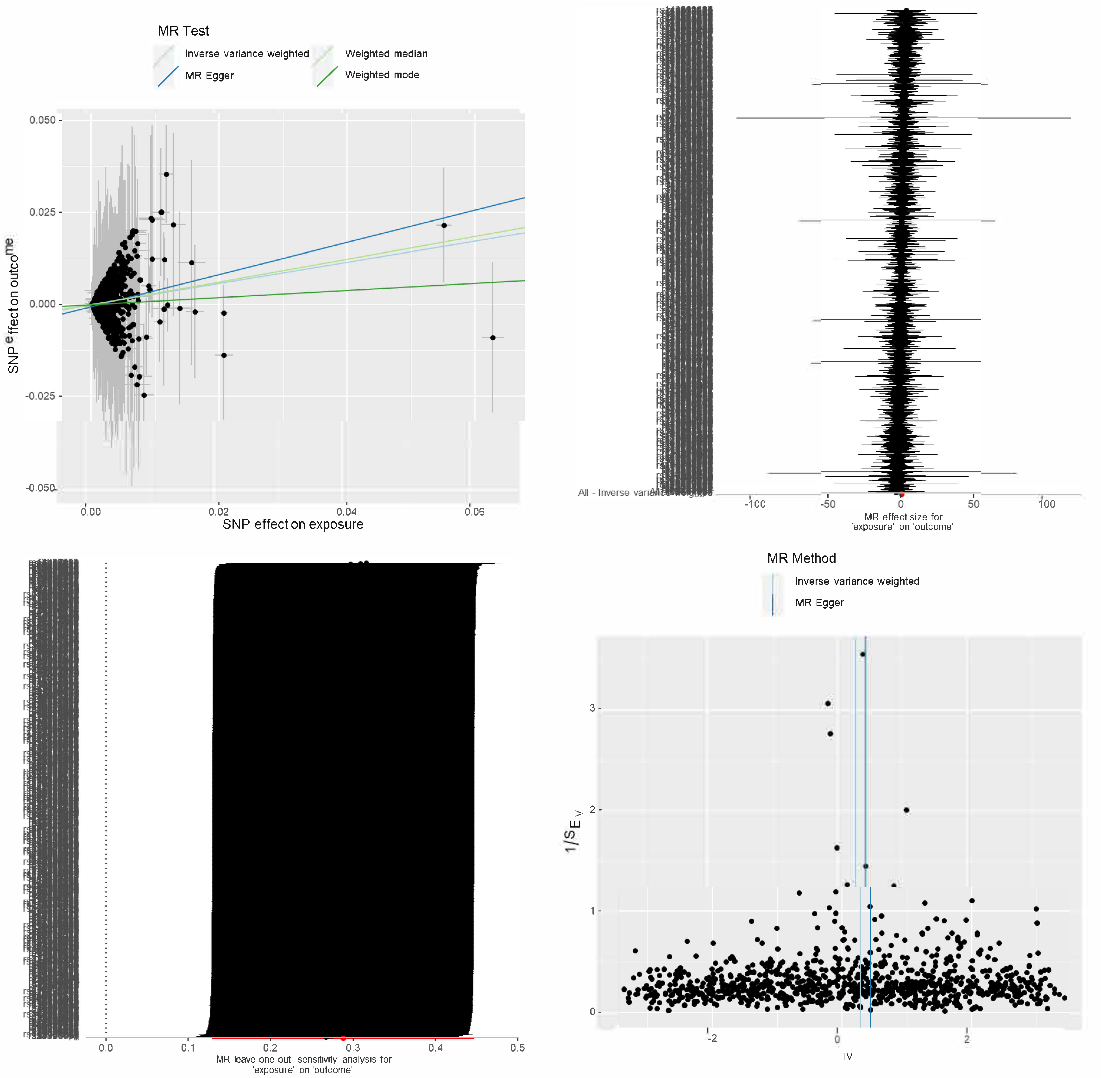

**Figure S3.** GSMR of causal association between eGFR^SCr^ and LVCO.

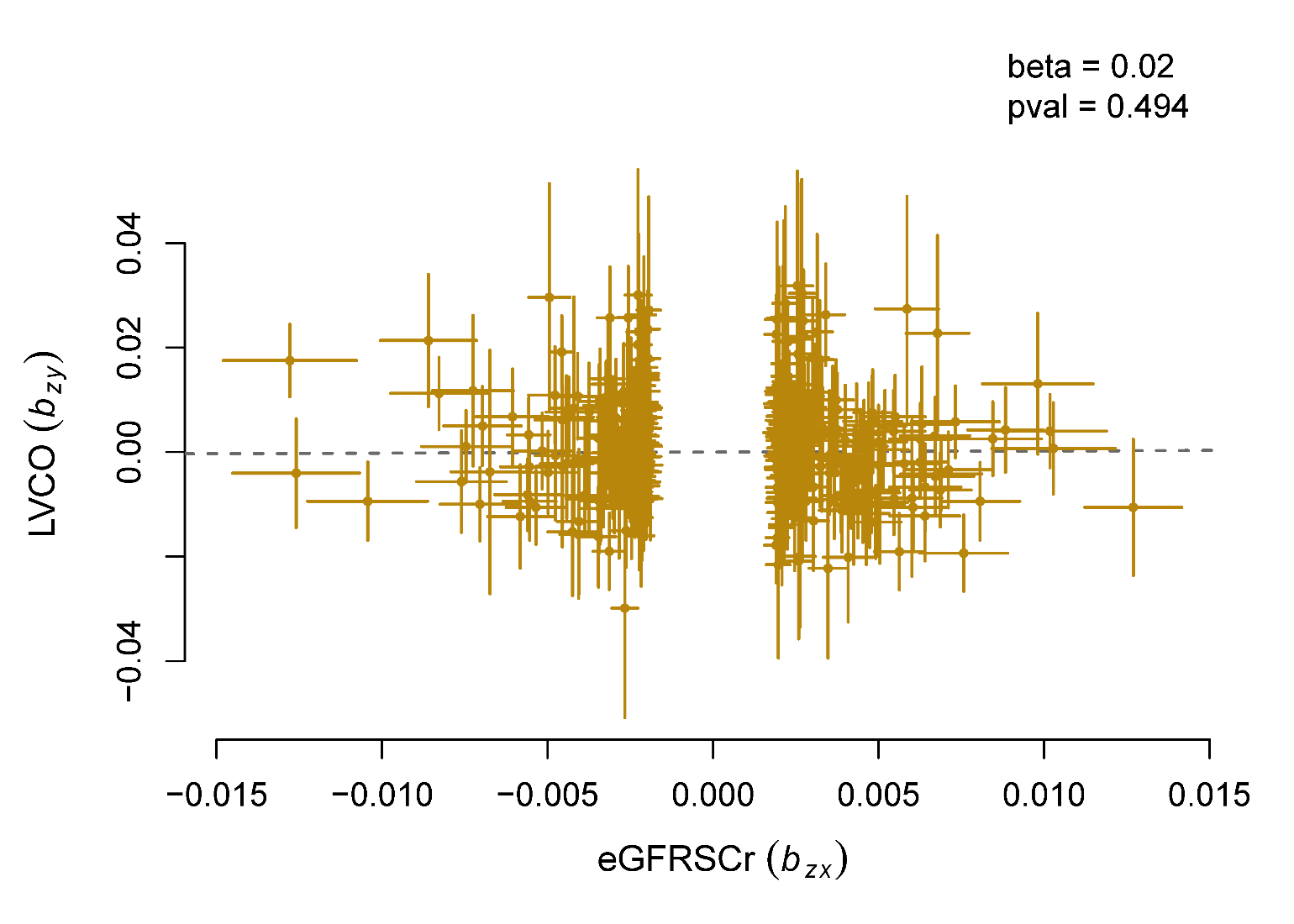

**Figure S4.** Sensitivity analysis of causal association between eGFR^SCr^ and LVCO.

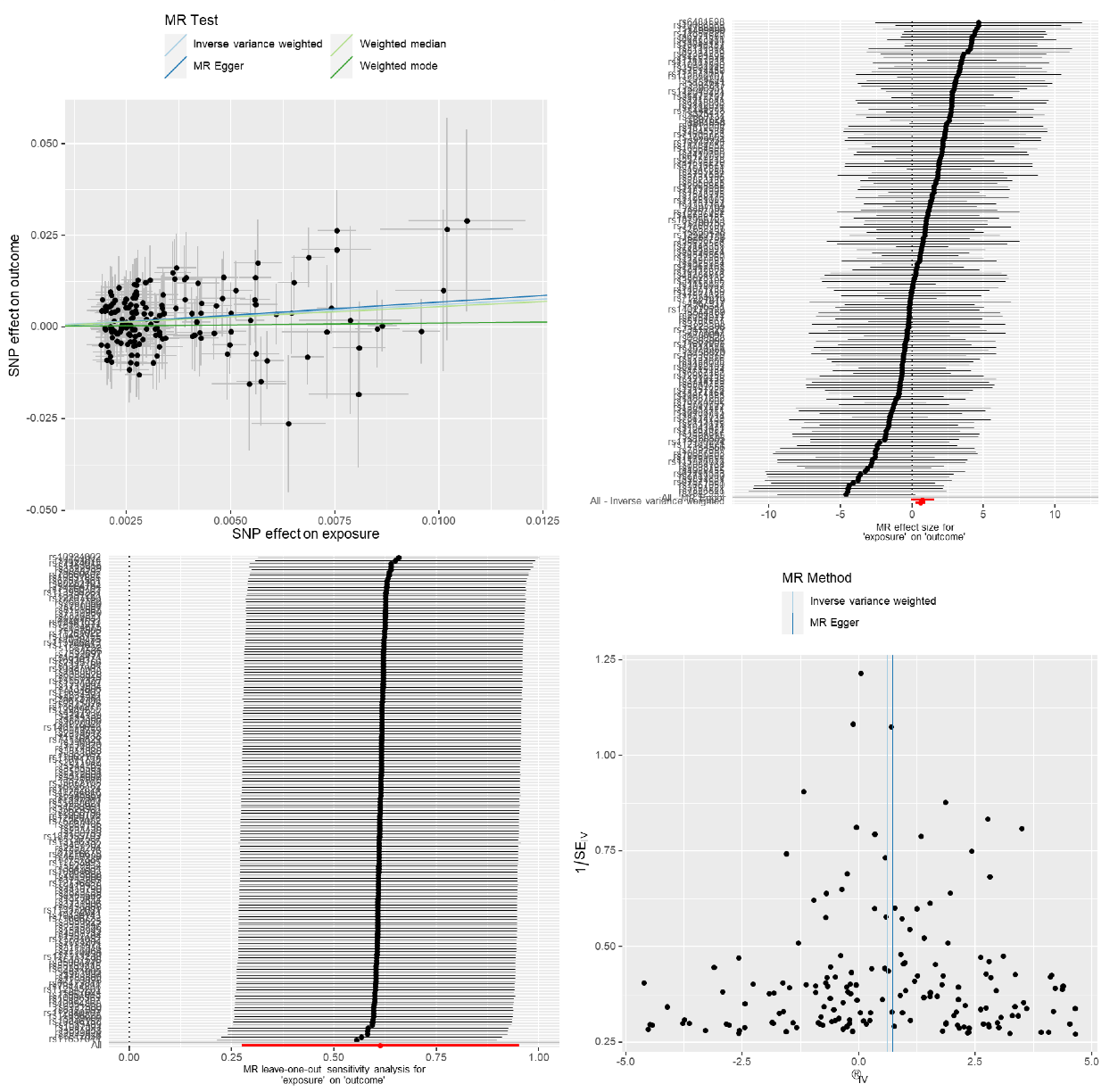

**Figure S5.** GSMR of causal association between uACR and LVM.

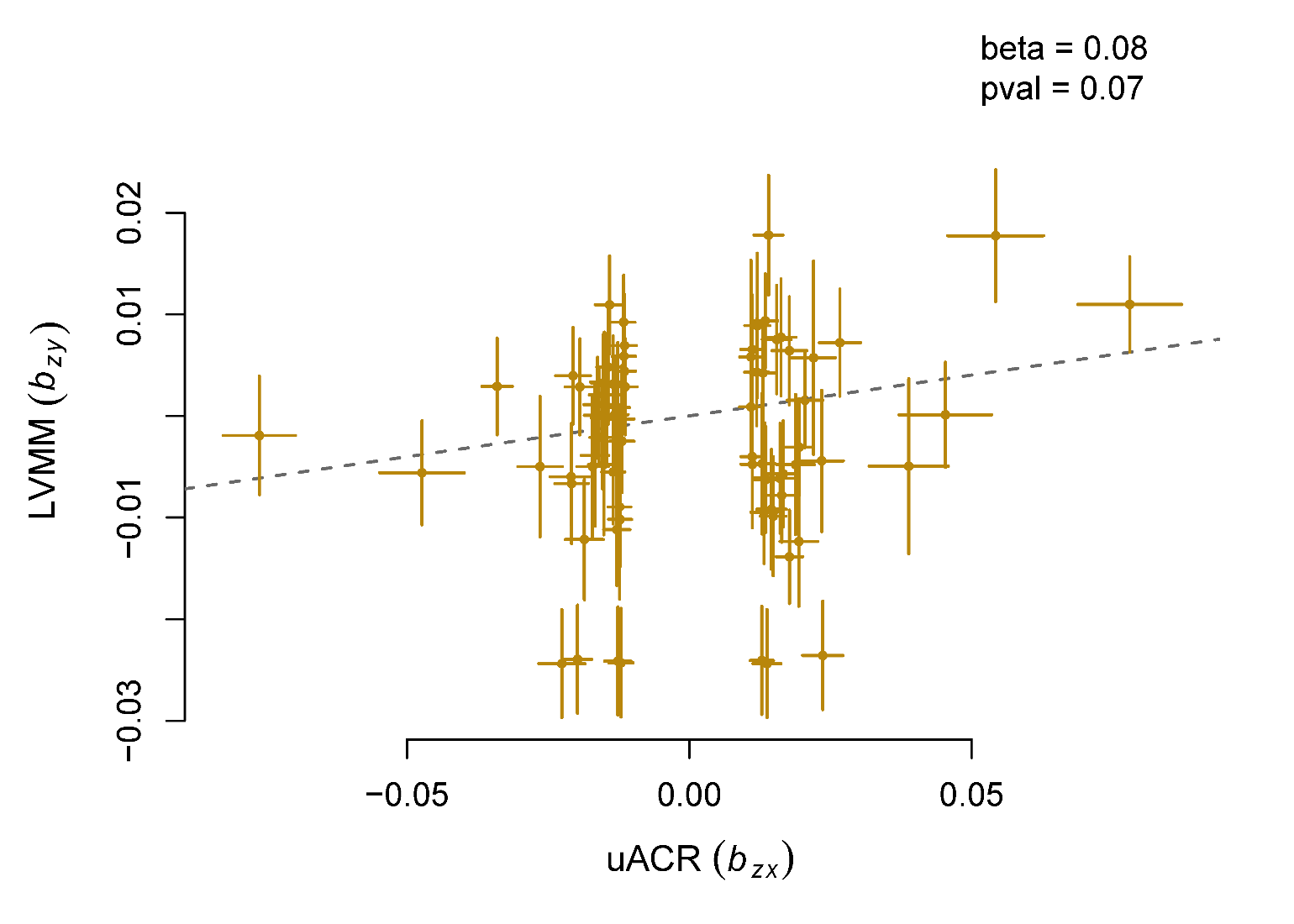

**Figure S6.** Sensitivity analysis of causal association between uACR and LVM.

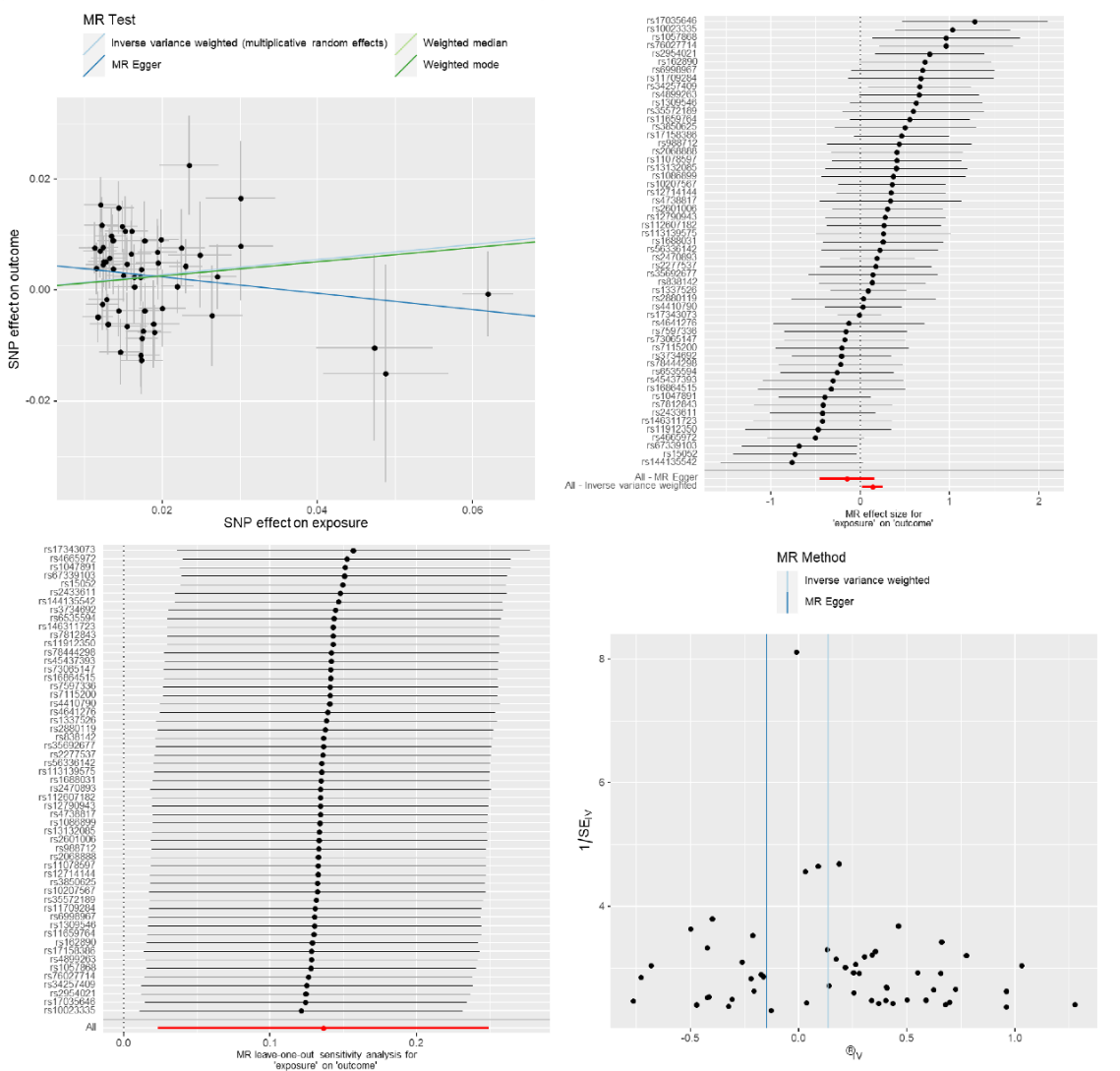

**Figure S7. Local genetic covariance estimates.**

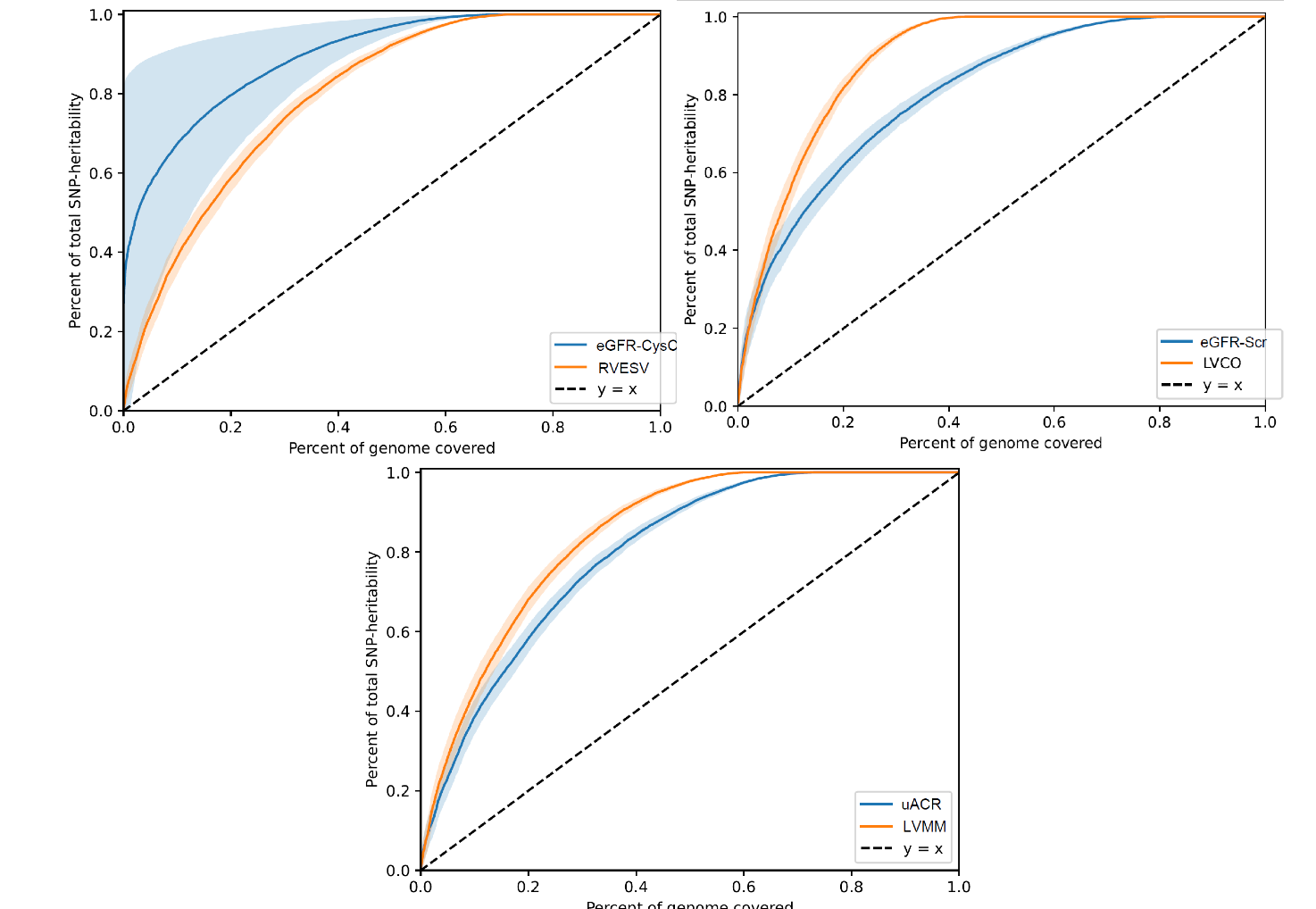

**Figure S8.** Local genetic correlation between eGFR^Scr^ and LVCO.

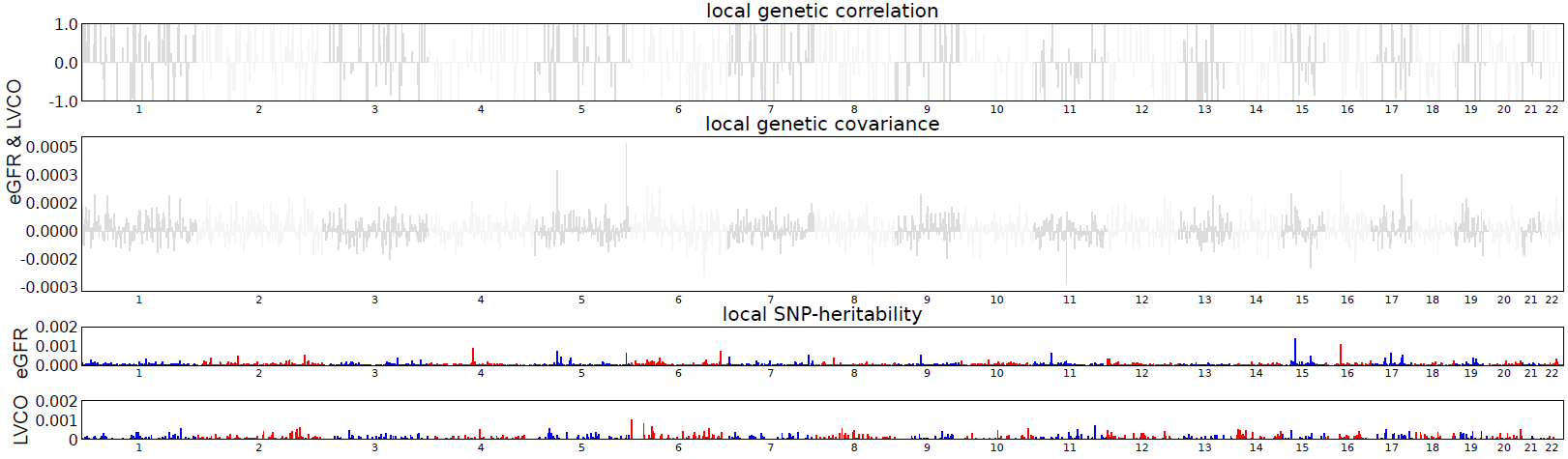

**Figure S9.** Local genetic correlation between uACR and LVM.

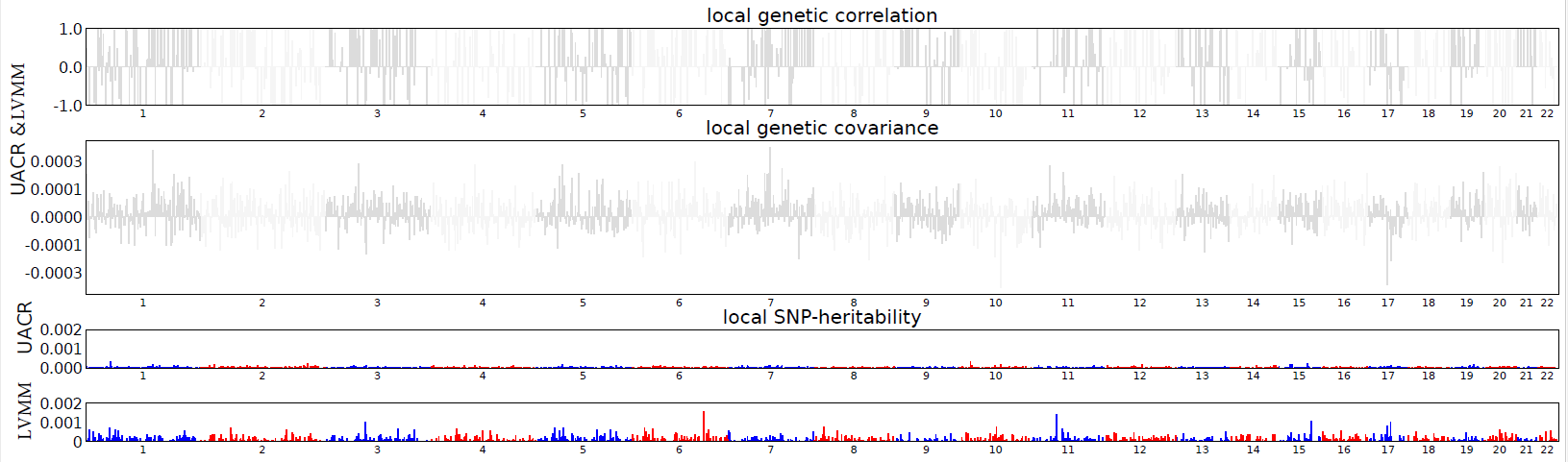

**Figure S10.** Colocalization analysis of eGFR^CysC^ and RVESV.

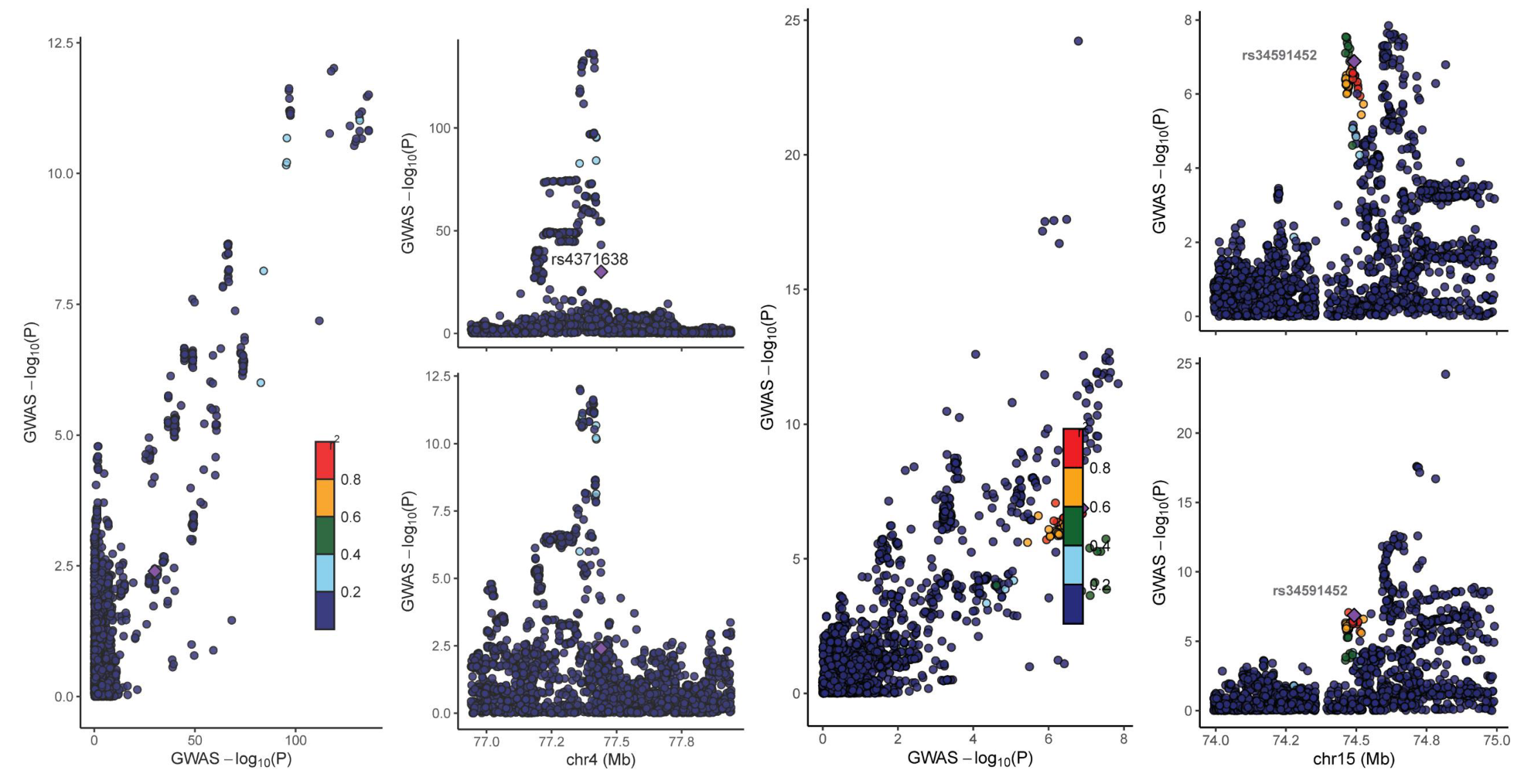

**Figure S11.** Study design of Mendelian Randomization Analysis.

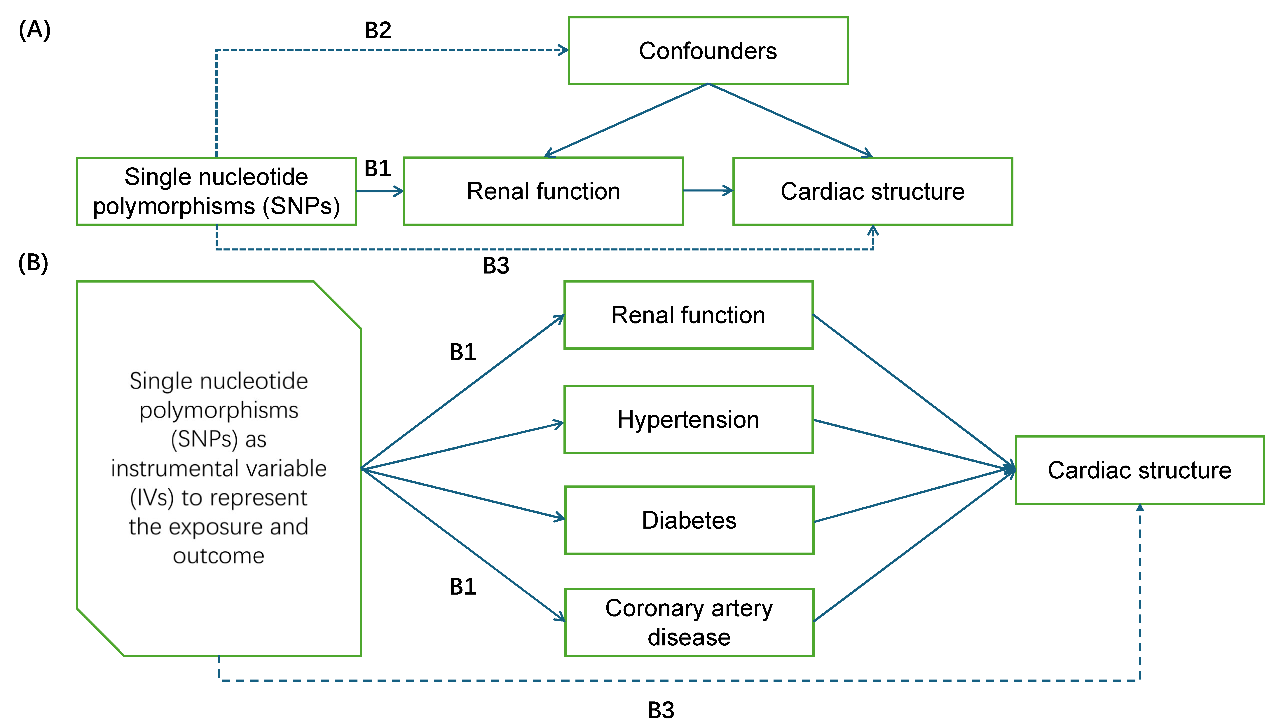

(A) Study design of univariateMendelian Randomization Analysis (B) Study design of Multivariable Mendelian Randomization Analysis; First, there must be a strong association between the IVs and the exposure variable (B1). Second, the IVs should be independent of known confounders (B2). Finally, the genetic variants should affect the outcome exclusively through the exposure pathway (B3) ensuring no direct involvement in the outcome

**Figure S12.** Pathway enrichment analysis of shared SNPs of eGFR and RVESV.

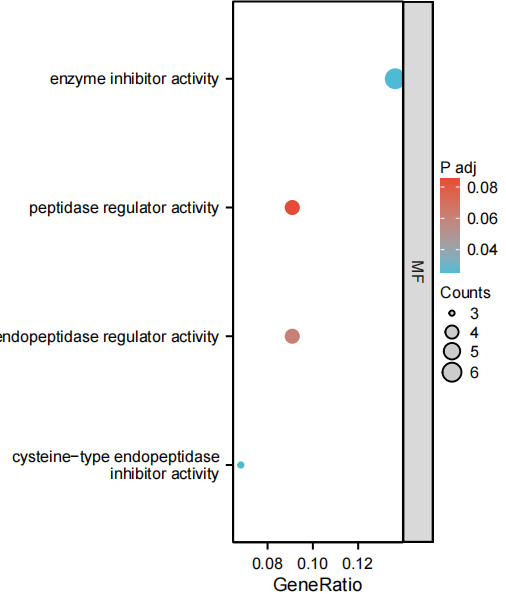
