## Supplementary material for "Mapping renal impairment and cardiac structure and function: a comprehensive analysis of prospective cohort study, Mendelian randomization and shared genetic etiology": checklist

**STROBE-MR checklist of recommended items to address in reports of Mendelian randomization studies**^1^ ^2^

| **Item No.** | **Section** | **Checklist item** | **Page No.** | **Relevant text from manuscript** |
| --- | --- | --- | --- | --- |
| 1 | **TITLE and ABSTRACT** | Indicate Mendelian randomization (MR) as the study’s design in the title and/or the abstract if that is a main purpose of the study | 1 | Mapping renal impairment and cardiac structure and function: a comprehensive analysis of prospective cohort study, Mendelian randomization and shared genetic etiology  Design: A comprehensive analysis of prospective cohort study, Mendelian randomization and shared genetic etiology. |
|  | **INTRODUCTION** |  |  |  |
| 2 | **Background** | Explain the scientific background and rationale for the reported study. What is the exposure? Is a potential causal relationship between exposure and outcome plausible? Justify why MR is a helpful method to address the study question | 3 | The UK Biobank (UKB), one of the largest multimodal studies, provides high-quality standardized CMR examinations and dense genotype data. This resource enables the establishment of genome-wide association studies (GWAS) profiles for CMR phenotypes, which can aid in evaluating genetic susceptibility to disease, developing new drug targets, and offering genetic insights into the kidney-heart connection (10). |
| 3 | **Objectives** | State specific objectives clearly, including pre-specified causal hypotheses (if any). State that MR is a method that, under specific assumptions, intends to estimate causal effects | 4 | Hence, this study aims to examine: 1) observational associations between renal function (CKD, eGFR [serum creatinine, cystatin C], uACR) and cardiac morphology, function, and geometry; 2) genetic associations between renal function and CMR measurements using linkage disequilibrium score regression (LDSC) and Mendelian randomization (MR); and 3) shared risk single nucleotide polymorphisms (SNPs) by employing Heritability Estimation from Summary Statistics (ρ-HESS), cross-trait meta-analyses, and colocalization. |
|  | **METHODS** |  |  |  |
| 4 | **Study design and data sources** | Present key elements of the study design early in the article. Consider including a table listing sources of data for all phases of the study. For each data source contributing to the analysis, describe the following: |  |  |
|  | a) | Setting: Describe the study design and the underlying population, if possible. Describe the setting, locations, and relevant dates, including periods of recruitment, exposure, follow-up, and data collection, when available. | NA |  |
|  | b) | Participants: Give the eligibility criteria, and the sources and methods of selection of participants. Report the sample size, and whether any power or sample size calculations were carried out prior to the main analysis | 6 | To explore potential causality and directionality between kidney function and CMR traits, we conducted MR analyses using large-scale GWAS datasets. Summary-level data for eGFR based on CysC (eGFRCysC) and uACR were obtained from a meta-analysis of European-ancestry participants from the CKDGen Consortium and UKB (15). The meta-analysis of the GWAS of eGFR based on creatine (eGFRSCr) was from CKDGen Consortium (16). The meta-analysis for eGFR based on creatinine (eGFRSCr) was sourced from CKDGen. GWAS summary statistics for CKD comprised 10,039 cases and 396,706 controls from the FinnGen study, identified via ICD codes. Summary-level data related to cardiac structure and function were obtained from the UKB meta-analysis (10). |
|  | c) | Describe measurement, quality control and selection of genetic variants | 6 |  |
|  | d) | For each exposure, outcome, and other relevant variables, describe methods of assessment and diagnostic criteria for diseases | 6 |  |
|  | e) | Provide details of ethics committee approval and participant informed consent, if relevant | 6 |  |
| 5 | **Assumptions** | Explicitly state the three core IV assumptions for the main analysis (relevance, independence and exclusion restriction) as well assumptions for any additional or sensitivity analysis | Supplement Method |  |
| 6 | **Statistical methods: main analysis** | Describe statistical methods and statistics used |  |  |
|  | a) | Describe how quantitative variables were handled in the analyses (i.e., scale, units, model) | NA |  |
|  | b) | Describe how genetic variants were handled in the analyses and, if applicable, how their weights were selected | 7 | We identified causal relationships using genetic instruments that reflect different aspects of renal pathophysiology: 1) index SNPs for eGFRCysC (Table S1), 2) eGFRSCr (Table S2), 3) CKD based on ICD (Table S3), and 4) uACR (Table S4). Genetic instruments were selected based on a GWAS-correlated P-value of 5 × 10-8, LD r² < 0.001, and being located within 1 MB of the index variant. Quality control filters applied included: MAF > 0.01; an F-statistic for each instrument (F = [(N-k-1)/k] * [R²/(1-R²)]) of < 10 indicating weak instruments; the Steiger filtering test to avoid reverse causality; and harmonization of exposure and outcome data. |
|  | c) | Describe the MR estimator (e.g. two-stage least squares, Wald ratio) and related statistics. Detail the included covariates and, in case of two-sample MR, whether the same covariate set was used for adjustment in the two samples | 7 | For each analysis, we used the inverse variance-weighted (IVW) MR method while accounting for random effects. |
|  | d) | Explain how missing data were addressed | NA |  |
|  | e) | If applicable, indicate how multiple testing was addressed | 7 | With 112 MR estimates (4 × 28), a Bonferroni-corrected P-value threshold was set at 0.05/112 (4.46 × 10-4), with P < 0.05 deemed nominally significant. |
| 7 | **Assessment of assumptions** | Describe any methods or prior knowledge used to assess the assumptions or justify their validity |  |  |
| 8 | **Sensitivity analyses and additional analyses** | Describe any sensitivity analyses or additional analyses performed (e.g. comparison of effect estimates from different approaches, independent replication, bias analytic techniques, validation of instruments, simulations) | 7 | We also reported the P-value for the intercept from MR Egger regression to check for horizontal pleiotropy. Sensitivity analyses included the weighted median method and MR-PRESSO to address measurement errors and selection bias. A combination of approaches provides robust evidence for potential causal relationships (16-18). |
| 9 | **Software and pre-registration** |  |  |  |
|  | a) | Name statistical software and package(s), including version and settings used | 7 | All analyses were performed using the TwoSampleMR (version 0.4.25) and MRPRESSO (version 1.0) packages in R (version 4.3). With 112 MR estimates (4 × 28), a Bonferroni-corrected P-value threshold was set at 0.05/112 (4.46 × 10-4), with P < 0.05 deemed nominally significant. |
|  | b) | State whether the study protocol and details were pre-registered (as well as when and where) | NA |  |
|  | **RESULTS** |  |  |  |
| 10 | **Descriptive data** |  |  |  |
|  | a) | Report the numbers of individuals at each stage of included studies and reasons for exclusion. Consider use of a flow diagram |  |  |
|  | b) | Report summary statistics for phenotypic exposure(s), outcome(s), and other relevant variables (e.g. means, SDs, proportions) | NA |  |
|  | c) | If the data sources include meta-analyses of previous studies, provide the assessments of heterogeneity across these studies | NA |  |
|  | d) | For two-sample MR:  i.  Provide justification of the similarity of the genetic variant-exposure associations between the exposure and outcome samples  ii.  Provide information on the number of individuals who overlap between the exposure and outcome studies | 10 | It conforms to the two-sample Mendelian randomization analysis, with no significant overlap in sample size (<5%). |
| 11 | **Main results** |  |  |  |
|  | a) | Report the associations between genetic variant and exposure, and between genetic variant and outcome, preferably on an interpretable scale | 10 |  |
|  | b) | Report MR estimates of the relationship between exposure and outcome, and the measures of uncertainty from the MR analysis, on an interpretable scale, such as odds ratio or relative risk per SD difference | 10 | In Table 3, genetically instrumented reduced log-eGFRCysC was found to decrease RVESV (β = -0.43 mL, 95% CI: -0.53 mm to -0.33 mm, P = 1.60×10-5) and RVEDV (β = -0.34 mL, 95% CI: -0.43 mL to -0.25 mL, P =0.0004). |
|  | c) | If relevant, consider translating estimates of relative risk into absolute risk for a meaningful time period | NA |  |
|  | d) | Consider plots to visualize results (e.g. forest plot, scatterplot of associations between genetic variants and outcome versus between genetic variants and exposure) | Supplement Figure |  |
| 12 | **Assessment of assumptions** |  |  |  |
|  | a) | Report the assessment of the validity of the assumptions | 10 | We conducted a comprehensive MR study from genetically predicted eGFR, CKD, and uACR on 28 cardiac structure and function, and identified 20 significant or nominal significant outcomes influenced by kidney function decline. The fixed-effects IVW method was used as our principal MR analytic approach. |
|  | b) | Report any additional statistics (e.g., assessments of heterogeneity across genetic variants, such as *I^2^*, Q statistic or E-value) | 10 | Heterogeneity was not observed with a Cochran Q-derived P value > 0.05 and the P value for the MR-Egger intercept is > 0.05 (Table S9). |
| 13 | **Sensitivity analyses and additional analyses** |  |  |  |
|  | a) | Report any sensitivity analyses to assess the robustness of the main results to violations of the assumptions | 10 | Heterogeneity was not observed with a Cochran Q-derived P value > 0.05 and the P value for the MR-Egger intercept is > 0.05 (Table S9). Similarly, Weighted median, MR-PRESSO, c-MRL, GSMR method also showed that reduced log-eGFRCysC was significantly correlated with an increased risk of RVESV and RVEDV (P <0.05) (Table S10 and Figure S1). |
|  | b) | Report results from other sensitivity analyses or additional analyses |  |  |
|  | c) | Report any assessment of direction of causal relationship (e.g., bidirectional MR) | 10 | Reverse causation was not observed in genetically determined reduced l log-eGFRCysC with RVESV and RVEDV (Table S10). |
|  | d) | When relevant, report and compare with estimates from non-MR analyses | 2 | Design: A comprehensive analysis of prospective cohort study, Mendelian randomization and shared genetic etiology. |
|  | e) | Consider additional plots to visualize results (e.g., leave-one-out analyses) | Supplement Figure |  |
|  | **DISCUSSION** |  |  |  |
| 14 | **Key results** | Summarize key results with reference to study objectives | 14 | This study represents the first extensive investigation into the potential causal effects of early kidney impairment on a broad spectrum of cardiac structure and function assessed through CMR traits. The observational and genetic data presented emphasize the importance of declining kidney function as a critical risk factor for alterations in cardiac structure and function. Notably, genetically instrumented early reductions in eGFR were significantly associated with decreased biventricular volume parameters and LVCO. Furthermore, we identified 20 novel shared loci associated with eGFRCysC decline and reduced RVESV, with two loci (rs4371638 at SHROOM3 and rs34591452 at STRA6) demonstrating strong evidence of colocalization. |
| 15 | **Limitations** | Discuss limitations of the study, taking into account the validity of the IV assumptions, other sources of potential bias, and imprecision. Discuss both direction and magnitude of any potential bias and any efforts to address them | 18 | Several limitations should be acknowledged. Firstly, there existed considerable variation in the estimations of causal effects derived from various variants (eGFR, CKD, uACR, Tables S1-S4). |
| 16 | **Interpretation** |  |  |  |
|  | a) | Meaning: Give a cautious overall interpretation of results in the context of their limitations and in comparison with other studies | 15 | Many large observational cohort studies, particularly those focused on CKD, have reported that declining renal function correlates with worsening diastolic function, a phenomenon confirmed by animal studies (4, 21-24). For instance, Buckley et al. reported that lower eGFR was linked to significant increases in left ventricular end-diastolic volume index and deteriorating diastolic measures among adults free of HF at baseline, while uACR did not correlate with changes in cardiac structure or function (23). Conversely, Cai et al. found that more advanced CKD at baseline correlated with increases in LVM and volume, as well as greater diastolic dysfunction (24). However, these results should be interpreted cautiously, as they may not entirely apply to community populations. Although up to 40% of CKD patients exhibit left ventricular hypertrophy (25), of interest, several studies report no independent association between lower kidney function and LVM. This discrepancy may arise from restrictive enrollment criteria in those studies (26, 27) , limiting the variability of cardiac structure. Some research suggests a positive relationship between high GFR and LV hypertrophy, implying a potential link between hyperfiltration and LV hypertrophy (28, 29). In mildly reduced eGFR stages, traditional risk factors such as hypertension may explain associations with LVM. Indeed, in community-based cohorts, the connection between low eGFR and LV hypertrophy was only significant before controlling for confounders (30). Consequently, the relationship between eGFR and left ventricular function and structure, especially within normal or mildly reduced ranges, may be complex. Buckley et al. found kidney function decline was not significantly associated with right ventricular fractional area change (23), although the limitations of cardiac ultrasound do not detract from the importance of assessing the right heart structure in relation to kidney function decline (31). Our study, conducted within a healthier community cohort, suggests that early cardiac changes relating to renal function decline differ in healthy populations. Mild renal dysfunction may correlate more closely with reduced stroke volume and decreased effective circulating blood volume. |
|  | b) | Mechanism: Discuss underlying biological mechanisms that could drive a potential causal relationship between the investigated exposure and the outcome, and whether the gene-environment equivalence assumption is reasonable. Use causal language carefully, clarifying that IV estimates may provide causal effects only under certain assumptions | 16 | The underlying mechanisms may include several factors. In the UKB population, 59.1% of participants had normal renal function (eGFR > 90 mL/min/1.73m²), while 39.3% had mild renal dysfunction (60 ≤ eGFR < 90 mL/min/1.73m²) and 2.7% had moderate to severe dysfunction (eGFR < 60 mL/min/1.73m²). In contrast, an Atherosclerosis Risk in Communities study indicated that 36.5% had reduced eGFR < 60 mL/min/1.73m² (4). |
|  | c) | Clinical relevance: Discuss whether the results have clinical or public policy relevance, and to what extent they inform effect sizes of possible interventions | 17 | In our study, we screened 9 SNPs with pleiotropic effects in the cross-trait meta-analyses and ρ-HESS of eGFRCysC and RVESV. Previous studies showed that SHROOM3 (rs4371638) associated with CKD showed associations with both the high levels of oxidatively damaged DNA and genomic instability (36, 37). SEMA7A(rs11854025) encodes a member of the semaphorin family of proteins. The encoded protein is found on activated lymphocytes and erythrocytes and may be involved in immunomodulatory and neuronal processes. SEMA7A is increased in patients with acute aortic dissection and is a Novel Biomarker in Kidney Renal Clear Cell Carcinoma (38, 39). NRG4 (rs10851885) is associated with imbalance in glucose metabolism and obesity, and significantly lower in patients with end-stage kidney disease (40-42). Notably, the independent shared SNP was rs2472297 (PCPASSOC = 2.86×10-33) located at CYP1A1. The cytochrome P450 proteins are monooxygenases which catalyze many reactions involved in drug metabolism and synthesis of cholesterol, steroids, and other lipids. Previous studies have linked CYP1A1 to ischemic stroke and coronary heart disease (43-45). These could be some potential targets for intervention. |
| 17 | **Generalizability** | Discuss the generalizability of the study results (a) to other populations, (b) across other exposure periods/timings, and (c) across other levels of exposure | 18 | In summary, our findings provide crucial insights for future research. Our logistic regression analyses established significant links between renal function deterioration and cardiac structure, demonstrating a notable association between reduced eGFR and both cardiac output and biventricular volume in the general population. We further confirmed a causal relationship between declining kidney function and alterations in cardiac structure and function, identifying key genes associated with early compensatory mechanisms. We recommend early CMR assessments for patients with mild renal insufficiency to accurately evaluate right ventricular function. |
|  | **OTHER INFORMATION** |  |  |  |
| 18 | **Funding** | Describe sources of funding and the role of funders in the present study and, if applicable, sources of funding for the databases and original study or studies on which the present study is based | 19 | This research was supported by grants from Guangdong Provincial Key Laboratory of Coronary Heart Disease Prevention (No. Y0120220151). |
| 19 | **Data and data sharing** | Provide the data used to perform all analyses or report where and how the data can be accessed, and reference these sources in the article. Provide the statistical code needed to reproduce the results in the article, or report whether the code is publicly accessible and if so, where | 20 | This study utilized data from the UK Biobank Resource (project number 99231). GWAS summary statistics for the 82 heart imaging traits are freely available for download from Zenodo: https://zenodo.org/record/7239166. Additional GWAS summary statistics and information from published projects of the CKDGen Consortium can be accessed at: https://ckdgen.imbi.uni-freiburg.de/. |
| 20 | **Conflicts of Interest** | All authors should declare all potential conflicts of interest | coi_disclosure.pdf |  |

This checklist is copyrighted by the Equator Network under the Creative Commons Attribution 3.0 Unported (CC BY 3.0) license.

1.	Skrivankova VW, Richmond RC, Woolf BAR, Yarmolinsky J, Davies NM, Swanson SA, et al. Strengthening the Reporting of Observational Studies in Epidemiology using Mendelian Randomization (STROBE-MR) Statement. JAMA. 2021;under review.

2.	Skrivankova VW, Richmond RC, Woolf BAR, Davies NM, Swanson SA, VanderWeele TJ, et al. Strengthening the Reporting of Observational Studies in Epidemiology using Mendelian Randomisation (STROBE-MR): Explanation and Elaboration. BMJ. 2021;375:n2233.
